# T cell responses to SARS-CoV-2 in people with and without neurologic symptoms of long COVID

**DOI:** 10.1101/2021.08.08.21261763

**Authors:** Lavanya Visvabharathy, Barbara A. Hanson, Zachary S. Orban, Patrick H. Lim, Nicole M. Palacio, Millenia Jimenez, Jeffrey R. Clark, Edith L. Graham, Eric M. Liotta, George Tachas, Pablo Penaloza-MacMaster, Igor J. Koralnik

## Abstract

Many people experiencing long COVID syndrome, or post-acute sequelae of SARS-CoV-2 infection (PASC), suffer from debilitating neurologic symptoms (Neuro-PASC). However, whether virus-specific adaptive immunity is affected in Neuro-PASC patients remains poorly understood. We report that Neuro-PASC patients exhibit distinct immunological signatures composed of elevated humoral and cellular responses toward SARS-CoV-2 Nucleocapsid protein at an average of 6 months post-infection compared to healthy COVID convalescents. Neuro-PASC patients also had enhanced virus-specific production of IL-6 from and diminished activation of CD8^+^ T cells. Furthermore, the severity of cognitive deficits or quality of life disturbances in Neuro-PASC patients were associated with a reduced diversity of effector molecule expression in T cells but elevated IFN-γ production to the C-terminal domain of Nucleocapsid protein. Proteomics analysis showed enhanced plasma immunoregulatory proteins and reduced pro-inflammatory and antiviral response proteins in Neuro-PASC patients compared with healthy COVID convalescents, which were also correlated with worse neurocognitive dysfunction. These data provide new insight into the pathogenesis of long COVID syndrome and a framework for the rational design of predictive biomarkers and therapeutic interventions.

**One Sentence Summary:** Adaptive immunity is altered in patients with neurologic manifestations of long COVID.

## Introduction

SARS-CoV-2 is the causative agent of a worldwide pandemic that was first identified in December, 2019. There have been more than 610 million cases and over 6 million deaths globally attributable to COVID-19 (1). Although highly effective vaccines are now used to prevent severe disease and death caused by SARS-CoV-2, long-term sequelae after infection have become an urgent medical concern.

SARS-CoV-2 infection can result in a wide spectrum of clinical manifestations ranging from asymptomatic infection to severe multi-organ dysfunction (2, 3), and predictive biomarkers to prognosticate either of these clinical outcomes are currently lacking. Globally, the estimated fatality rate following SARS-CoV-2 infection is approximately 2%, but not all patients recover to their baseline states (4). “Long COVID” affects an estimated 30% of people infected with SARS-CoV-2 and includes symptoms persisting more than 4 weeks after infection, termed “post-acute sequelae of SARS-CoV-2 infection” or PASC (5). According to the Centers for Disease Control and Prevention (CDC) and others, Neuro-PASC is clinically defined as new neurologic or neurocognitive symptoms persisting for more than 4 weeks after disease onset and is often not concomitant with diagnosis of acute infection (6, 7). Although the majority of people with SARS-CoV-2 infection experience mild disease not requiring hospitalization, more than half of these individuals have symptoms persisting more than 4 months after acute infection (8). This is similar to the frequency of neuropsychiatric symptoms reported by people infected Middle East Respiratory virus (MERS) and SARS-CoV-1 up to 3.5 years after acute infection (9), suggesting that SARS coronaviruses commonly cause long-term neurological sequelae. Similarly, recent studies on recovered COVID-19 patients showed significant cognitive deficits in attention, working memory, and emotional processing months after the resolution of acute infection (10).

T cell immunity is necessary for the host defense against SARS-CoV-2. In particular, CD4^+^ T cell responses directed against the Spike protein were found in 100% of COVID convalescents (11), and virus-specific T cell responses were sub-optimal or impaired in severely ill COVID patients (12). Autopsies of severe COVID patients found impaired germinal center formation linked to a defective T follicular helper cell response (13). Studies have also shown that CD8^+^ T cell depletion after SARS-CoV-2 infection of rhesus macaques impairs anamnestic immune protection after subsequent re-infection (14). Moreover, memory T cell responses can be detected in patients exposed to the closely related SARS-CoV-1 up to 4 years after virus exposure (15). Despite these studies showing a role for T cells in protecting against acute SARS-CoV-2 infection, the impact of T cell responses on PASC remains poorly understood. Therefore, we sought to determine how SARS-CoV-2-specific T cell responses contribute to the etiology and pathogenesis of PASC.

Here, we focus on a group of Neuro-PASC patients who mostly had mild acute disease but subsequently developed a substantial reduction in their quality of life, psychiatric, and cognitive parameters. Our data show four critical findings linking T cell responses with Neuro-PASC symptoms. Firstly, we show that Neuro-PASC patients exhibit decreased Spike-but increased Nucleocapsid- and Membrane-specific T cell responses compared with healthy COVID convalescents without persistent symptoms. Secondly, CD8^+^ memory T cells from Neuro-PASC patients produce substantially more IL-6 in response to Spike and Nucleocapsid peptides. Thirdly, the increased severity of cognitive deficits and decreased quality of life markers are positively correlated with IFN-γ production in response to Nucleocapsid antigens and altered effector molecule expression in memory T cells. Lastly, Neuro-PASC patients presented with higher levels of immunoregulatory proteins and lower levels of antiviral and T_h_1 inflammatory proteins in proteomics analysis. Together, these data suggest wide-ranging immunological alterations in Neuro-PASC patients, with important implications for appropriate diagnostic, prevention, and treatment strategies.

## Results

### Clinical characteristics of Neuro-PASC patients and control participants

We enrolled a total of 168 participants, including 143 prior to SARS-CoV-2 vaccination and 25 participants post-vaccination recruited from the Neuro-COVID-19 outpatient clinic at Northwestern Memorial Hospital or from the surrounding Chicago area. These included 91 Neuro-PASC patients (“NP”; confirmed RT-PCR+ or anti-SARS-CoV-2 Spike IgG+) meeting Infectious Disease Society of America clinical criteria for COVID-19 starting after February 2020 and had neurologic symptoms lasting at least 6 weeks post-infection, as previously reported (16). Among those, 66 (80.6%) were never hospitalized for pneumonia or hypoxia and had mild disease. We additionally recruited 43 COVID convalescents without lasting symptoms (“CC”; RT-PCR+ or seropositive for anti-SARS-CoV-2 Spike RBD IgG); and 34 healthy controls who were RT-PCR- and seronegative for SARS-CoV-2 Spike-IgG (“HC”; study design in Fig. 1A).

**Figure 1:**
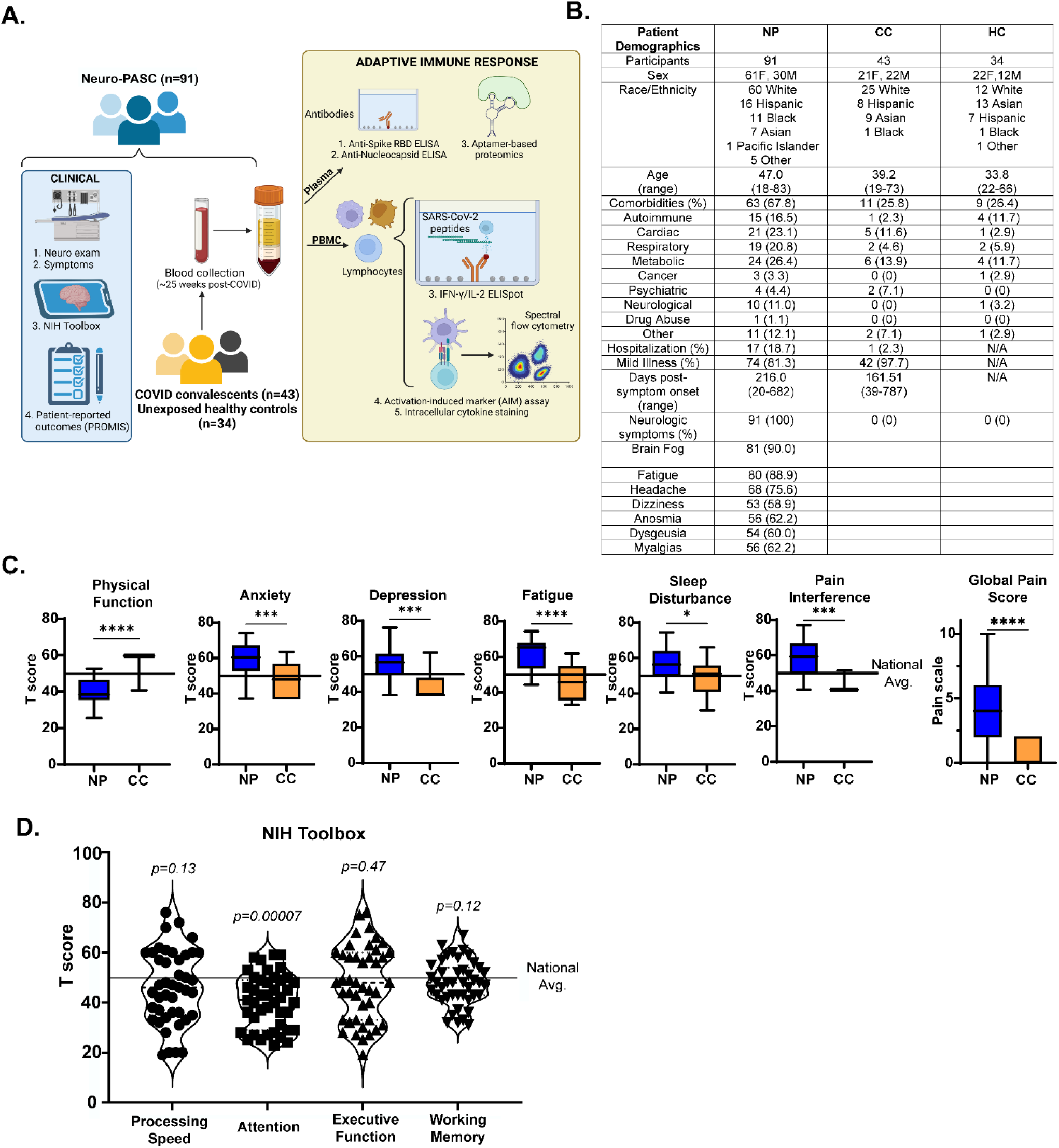
Study design and clinical data. A.) Study design, including clinical and immunological data collection. B.) Demographic table for Neuro-PASC, COVID convalescent, and healthy control subjects. C.) PROMIS-57 patient-reported survey T scores for Neuro-PASC patients (n=36) and COVID convalescents (n=13). D.) NIH Toolbox cognitive T scores for Neuro-PASC patients (n=55). Horizontal black line represents the U.S. national average T score of 50; *p* values relative to demographic-matched US national average by one sample Wilcoxon signed rank test. *p<0.05, ***p<0.005, ****p<0.0001 by two-tailed Student’s t test.

Neuro-PASC patients displayed a constellation of neurological symptoms similar to those previously reported (17) such as headache, fatigue, brain fog, and myalgia (Fig. 1B). Results from the patient reported outcomes information system (PROMIS-57) survey (18) showed that Neuro-PASC patients scored significantly lower on physical function and higher on anxiety, depression, pain and other quality of life metrics compared with COVID convalescents or the national average (Fig. 1C). NIH toolbox tests administered to assess cognitive function (19) in Neuro-PASC patients also showed significantly lower T scores in the attention module, indicative of cognitive dysfunction relative to a demographic-matched population (Fig. 1D).

### Neuro-PASC T cell responses to SARS-CoV-2 structural proteins

To determine the specificity of T cell responses to SARS-CoV-2 in Neuro-PASC and COVID convalescent groups, we performed cytokine ELISPOT. Bulk peripheral blood mononuclear cells (PBMC) from each subject were stimulated with overlapping peptides from the Spike (S), Nucleocapsid (N), or Membrane (M) structural proteins of SARS-CoV-2 (Fig. S1). IFN-γ ^+^ and IL-2 ^+^ T cell responses to S peptides were similar between Neuro-PASC patients and COVID convalescents (Fig. 2A left panel, S2A). However, Neuro-PASC patients exhibited higher IFN-γ ^+^ T cell responses against N and M peptides (Fig. 2A, right panels) compared with COVID convalescents. Further experiments dividing Nucleocapsid peptides into 3 pools (Fig. 2B) pinpointed the increased T cell reactivity in Neuro-PASC patients to the C-terminal region of the protein (Fig. 2C), particularly in amino acids 309-402 (Fig. 2D). T cell receptor (TCR sequencing) was then performed on a subset of study participants. The top N3 region-specific TCR clone was more highly expanded in Neuro-PASC patients than COVID convalescents (Fig. 2E), consistent with IFN-γ responses in Fig. 2C. Antibody titers against the Spike receptor-binding domain (RBD) did not differ between Neuro-PASC and COVID convalescent groups (Fig. 2F). No differences in antibody titers against the irrelevant Haemagglutinin protein from Influenza virus were found between groups (Fig. 2G), demonstrating immune responses were SARS-CoV-2-specific.

**Figure 2:**
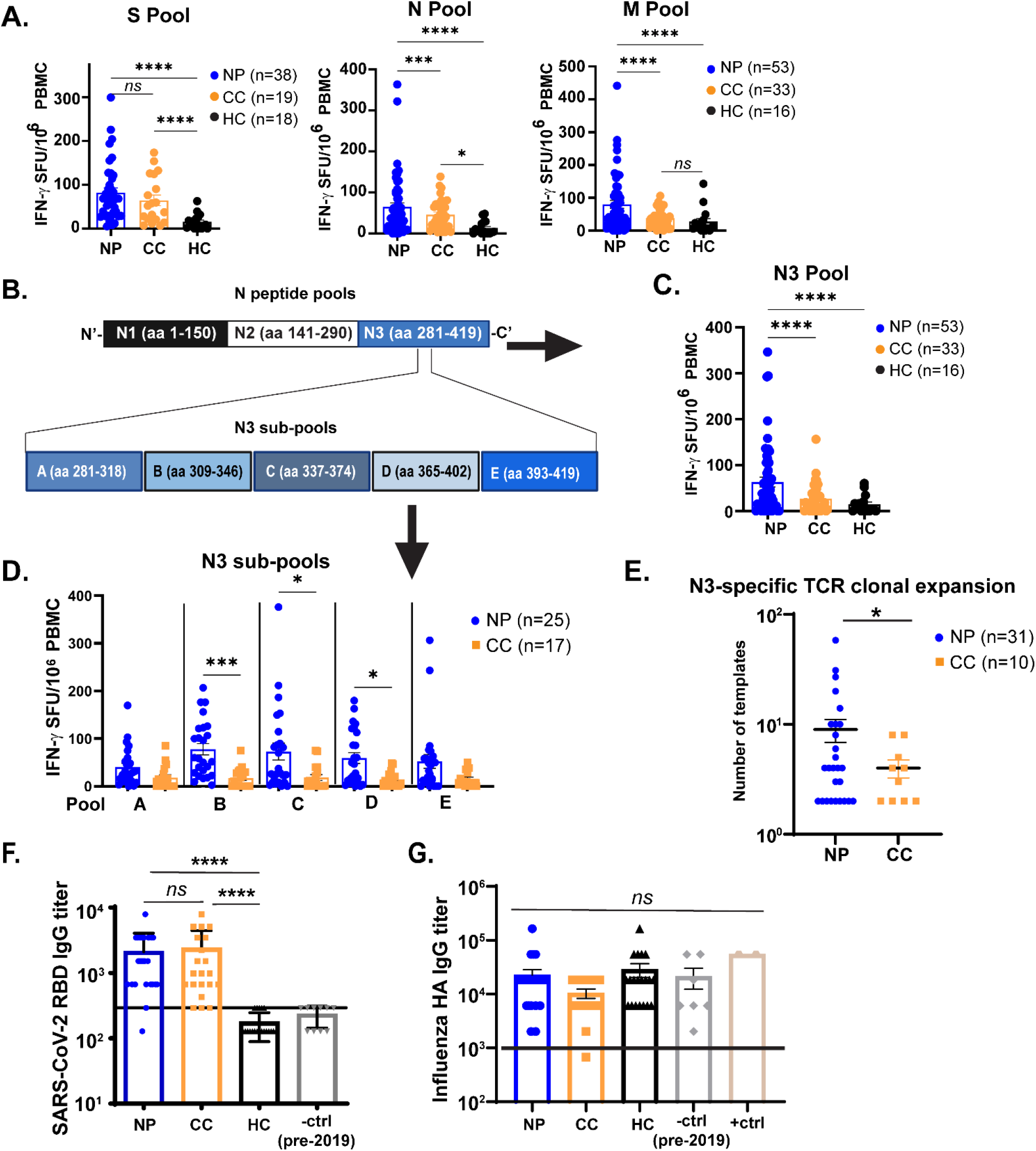
T cells from Neuro-PASC patients have elevated responses to SARS-CoV-2 Nucleocapsid and Membrane proteins compared to COVID convalescents. A.) Unvaccinated Neuro-PASC patients and healthy COVID convalescents display similar IFN-γ responses to SARS-CoV-2 S peptides, but Neuro-PASC patients have enhanced N- and M peptide-specific responses. B.) Diagram of sub pools derived from N protein (N1-N3, top) and further subdivision of the N3 peptide pools into 5 sub pools (A-E, bottom). C.) Neuro-PASC T cells have enhanced IFN-γ responses to the C-terminal region of the N protein (N3) compared to control groups. D.) Neuro-PASC T cells are more highly reactive to certain regions of the C-terminal domain of N protein, particularly amino acids 309-346, of the SARS-CoV-2 Nucleocapsid protein compared with COVID convalescents. E.) Mapping of TCR sequences to SARS-CoV-2 Nucleocapsid peptide reveals enhanced N3-specific clonal expansion in Neuro-PASC patients. F.) Spike RBD antibody response quantification for all groups. G.) Influenza A hemagglutinin (HA) antibody responses for all groups. +ctrl = plasma from patients who received the Influenza vaccine within 3 weeks before sample collection. Vaccinated individuals were included in CC samples for Fig. 2A-D for N and M-specific responses, as SARS-CoV-2 vaccination does not impact T cell responses to viral proteins other than Spike. TCR antigen specificity was identified using the immuneCODE database from Adaptive Biotechnologies. Horizontal black line in F-G = limit of detection. Data representative of 10 experiments with all conditions plated in duplicate. *p<0.05, **p<0.01, ***p<0.005, ****p<0.0001 by one-way ANOVA with Tukey’s posttest (A,C, F) or t Test with Welch’s correction (D, E).

No significant differences were found in IL-2 and IL-17 cytokine responses between groups (Fig. S2B-F). Healthy controls exhibited some IL-2 production to N peptides likely caused by cross-reactivity with endemic coronaviruses (Fig. S2B) as suggested previously (20). Importantly, hospitalization prior to the development of Neuro-PASC did not affect the IFN-γ response to SARS-CoV-2 (Fig. S2G-H). Though post-hospitalized Neuro-PASC patients trended towards lower IFN-γ ^+^ T cell responses compared with non-hospitalized patients, these were statistically non-significant.

### Virus-specific activation of CD4^+^ Tfh cells in Neuro-PASC

Comparison of CD4^+^ T cell subsets from hospitalized and non-hospitalized COVID patients showed that severe disease was associated with elevated T follicular helper (Tfh) proportions relative to patients with mild disease (21). We thus determined whether Tfh activation (see gating scheme in Fig. S3C) could similarly differentiate Neuro-PASC patients from COVID convalescents. Immunophenotyping showed no differences between groups in total percentages of most T cell subsets, including Tfh cells, in the unstimulated condition (Fig. S4).

Therefore, we conducted functional assays to determine T cell reactivity. The activation-induced marker (AIM) assay measures cytokine-independent, antigen-specific, TCR-mediated T cell activation and has been previously used to detect SARS-CoV-2-specific CD4^+^ (CD137^+^CD134^+^) and CD8^+^ (CD69^+^CD137^+^) T cells (11). We used this method to investigate Tfh activation. N-specific CD134^+^CD137^+^ (AIM^+^) Tfh cells were significantly elevated in Neuro-PASC patients compared with COVID convalescents, while the opposite pattern was observed for S and M-specific Tfh cells (Fig. 3A-B). Consistent with these results, N-specific IgG titers were significantly elevated in Neuro-PASC patients (Fig. 3C). Neither N-specific Tfh cell activation nor anti-N antibody responses decreased with the time post-acute infection in Neuro-PASC patients (Fig. S5A-B).

**Figure 3:**
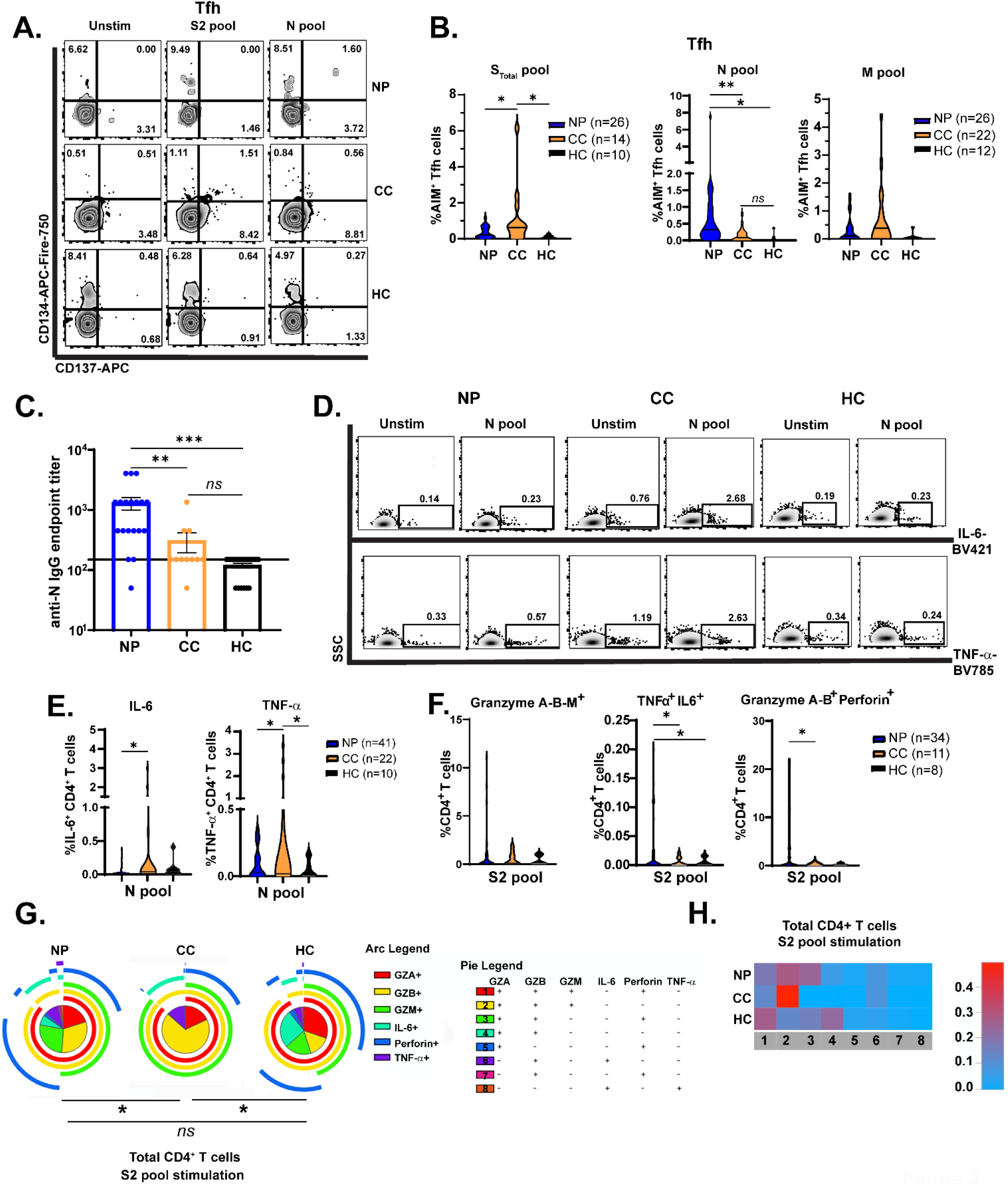
Virus-specific Tfh cell activation and CD4^+^ T cell effector functions differ between Neuro-PASC patients and COVID convalescents. A.) CD4^+^ Tfh cells from Neuro-PASC patients display reciprocal activation patterns to COVID convalescents in response to Spike and Nucleocapsid peptides. B.) Quantification of AIM^+^ Tfh cell activation to Spike, Nucleocapsid, and Membrane peptides. C.) Anti-SARS-CoV-2 Nucleocapsid IgG endpoint titers for Neuro-PASC, COVID convalescents, and healthy controls shown in B. D.) CD4^+^ T cells from Neuro-PASC patients express less IL-6 and TNF-α in response to N peptides compared with COVID convalescents. E.) Quantification of cytokine production from D. F.) CD4^+^ T cells from Neuro-PASC patients have enhanced polyfunctionality and granzyme production after Spike peptide stimulation compared with COVID convalescents. G.) Expression of cytolytic effector molecules in CD4^+^ T cells after S peptide stimulation. H.) Heatmap quantifying polyfunctionality in different categories of cytokine production between groups. All data for S pool obtained from unvaccinated individuals; data for N and M pools obtained from some vaccinated healthy COVID convalescent and healthy control subjects. Data combined from 6 independent experiments with the indicated n values. *p<0.05, **p<0.01, ***p<0.005 using one-way ANOVA with Bonferroni’s posttest (B, C); two-tailed Student’s t Test with Welch’s correction (E, F) or a Permutation test (G). All pie graphs are background subtracted (unstimulated condition; Fig. S7A).

### CD4^+^ T cell effector functions differ in Neuro-PASC patients vs. COVID convalescents

To probe the effector functions of virus-specific CD4^+^ T cells, we determined whether Neuro-PASC patients and COVID convalescents had altered patterns of cytokine production in response to viral antigens. We focused on T cell responses to N and S2 peptide pools because these antigens provoked maximal differences between Neuro-PASC patients and control groups. CD4^+^ T cells from Neuro-PASC patients expressed lower levels of IL-6 and TNF-α relative to COVID convalescents following stimulation with N peptides (Fig. 3D-E), and higher levels of TNF-α after S peptide stimulation (Fig. S6). No differences were observed in the unstimulated condition (Fig. S5C). CD4^+^ T cells can also produce cytolytic granules that help eliminate virus-infected cells (22) as in Influenza or HIV infection (23, 24). However, enhanced production of cytolytic granules from CD4^+^ T cells was also associated with disease severity in acutely infected COVID-19 patients (25). We therefore investigated cytolytic granule (granzyme A, B, M, and Perforin), IL-6, and TNF-α expression in CD4^+^ T cells following stimulation with SARS-CoV-2 peptides. Neuro-PASC patients had significant elevations in dual and triple cytokine- and cytolytic granule-producing CD4^+^ T cells after S pool stimulation, including in granzyme A/B^+^, TNF-α/IL-6^+^, and granzyme A/B-Perforin^+^ CD4^+^ T cells (Fig. 3F). S-specific CD4^+^ T cells from Neuro-PASC patients also retained polyfunctionality similar to healthy controls, while CD4^+^ T cells from COVID convalescents were limited to producing mostly granzymes (category 2 in yellow, Fig. 3G-H; unstimulated and N pool stimulation in Fig. S7A-B). These data suggest that cytotoxic responses to Spike protein in CD4^+^ T cells from Neuro-PASC patients are functionally distinct from those in COVID convalescents, and do not significantly differ from unexposed healthy controls.

### Attenuated CD8^+^ memory T cell activation in Neuro-PASC patients

CD8^+^ memory T cells are important for effective anti-viral immunity and can persist for several years after the related SARS-CoV-1 infection (26). However, little is known about memory CD8^+^ T cell function in Neuro-PASC. CD8^+^ T effector memory cells (TEM or TEMRA; gating strategy in Fig. S3A) are poised for rapid cytotoxic function upon antigen re-encounter. CD8^+^ TEM exhibited significant antigen-driven activation in COVID convalescents but not in Neuro-PASC patients (Fig. 4A-B). Total percentages of CD8^+^ TEMRA cells were also significantly elevated in Neuro PASC patients (Fig. 4C), but despite their increased numbers, these cells were less activated by S and N peptides compared with COVID convalescents (Fig. 4D-E). Similarly, virus-specific cytokine production differed in CD8^+^ T cell subsets between groups. S and N peptides provoked elevated IL-6 production on a per-cell basis in CD8^+^ TEM from Neuro-PASC patients compared to COVID convalescents (Fig. 4F-G), as determined by mean fluorescence intensity (MFI). Monocytes and neutrophils, innate immune cells that are among the main producers of IL-6 (27), also expressed significantly more IL-6 after stimulation with viral peptides in Neuro-PASC patients compared to control groups (Fig. S8A-B), possibly due to enhanced innate receptor stimulation (28, 29).

**Figure 4:**
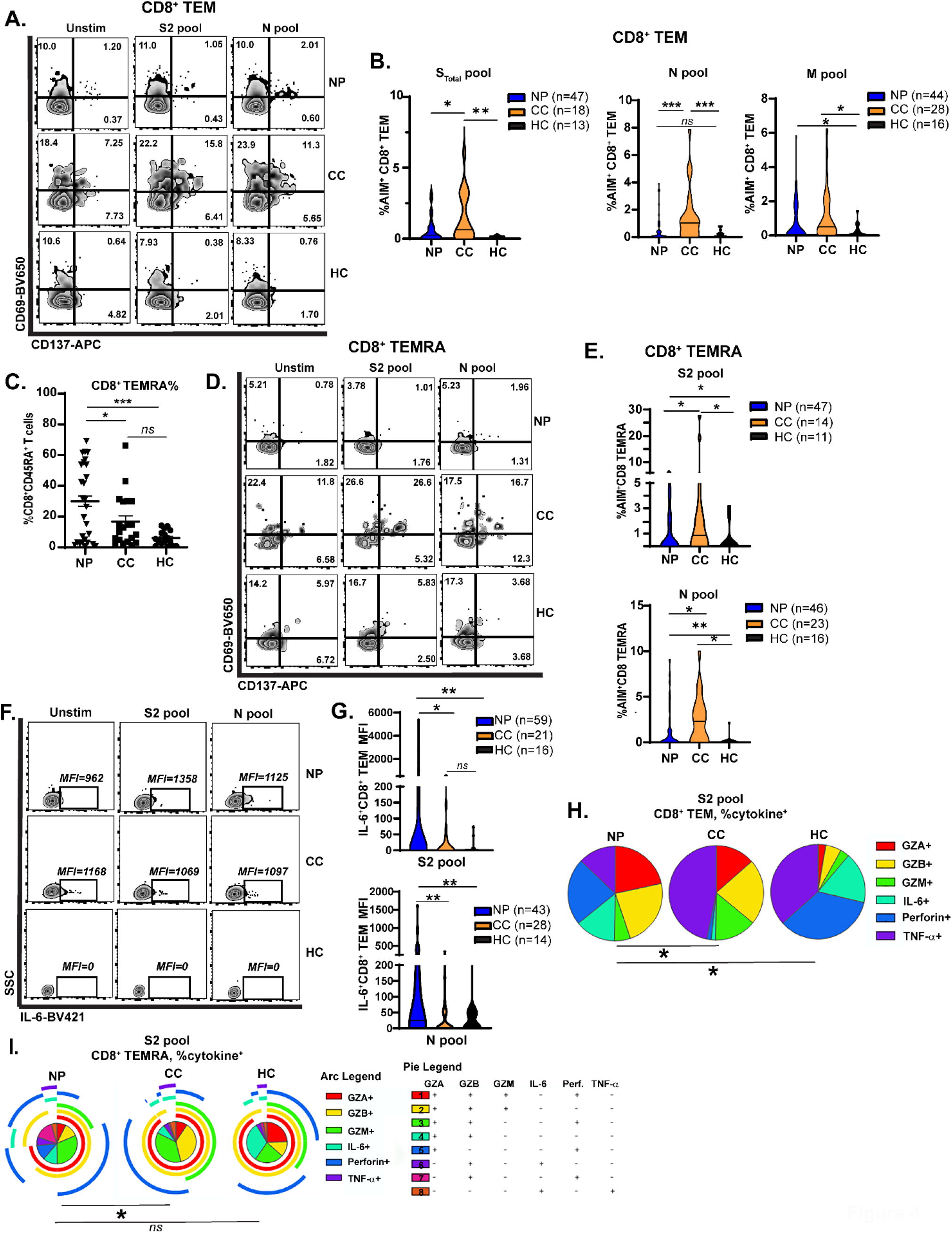
Altered CD8^+^ memory T cell activation and function in Neuro-PASC. A.) CD8^+^ TEM from Neuro-PASC patients show decreased activation after stimulation with viral peptides. B.) Quantification of CD8^+^ TEM cell activation after S, N, and M peptide stimulation. C.) CD8^+^ TEMRA cells accumulate significantly in PBMC from Neuro-PASC patients compared to control groups. D.) CD8^+^ TEMRA cells from Neuro-PASC patients are less activated by viral peptides compared with COVID convalescents. E.) Quantification of CD8^+^ TEMRA cell activation. F-G.) CD8^+^ TEM from Neuro-PASC patients have enhanced IL-6 production after S and N peptide stimulation compared to COVID convalescents on a per-cell basis as determined by mean fluorescence intensity (MFI). H.) Spike-specific CD8^+^ TEM express more TNF-α or granzyme B in COVID convalescents while those from Neuro-PASC patients are biased toward IL-6 and Perforin expression. All data for S pool obtained from unvaccinated individuals; data for N and M pools obtained from some vaccinated healthy COVID convalescent and healthy control subjects. Data combined from 5 independent experiments with the indicated n values. *p<0.05, **p<0.01, ***p<0.005 using two-tailed Student’s t test with Welch’s correction (B, C, E, G) or Permutation test (H). All pie graphs show data after subtracting background (unstimulated condition; Fig. S7 C, E).

Additionally, CD8^+^ T cells had differing patterns of cytolytic effector molecule production between groups. CD8^+^ TEM from Neuro-PASC patients exhibited elevated S-specific granzyme production compared with COVID convalescents or healthy controls (Fig. 4H; unstimulated and N pool stimulation in Fig. S7C-D). However, S peptides did not significantly alter CD8^+^ TEMRA effector functions in Neuro-PASC patients relative to unexposed healthy controls. In contrast, COVID convalescents produced more cytolytic granzymes and Perforin, and this functional change was consistent with the higher activation seen in Fig. 4E (Fig. 4I; unstimulated and N pool stimulation in Fig. S7E-F). Expression of the inhibitory receptor PD-1 did not differ in CD8^+^ TEM after stimulation with viral peptides in Neuro-PASC patients and COVID convalescents (Fig. S8C).

### Impaired cognition and decreased quality of life metrics correlate with distinct patterns of virus-specific T cell responses

We next probed whether within-group differences in adaptive immune responses correlated with clinical measures of symptom severity in Neuro-PASC. Poorer cognitive and anxiety scores were correlated with elevated IFN-γ-expressing T cells directed against the C-terminal domain of N protein (Fig. 5A). Spearman rank correlation analysis further demonstrated negative correlations between attention and executive function scores and IFN-γ responses to the C-terminal domain of N protein as well as RBD-specific antibody responses (Fig. 5B), among other parameters. To determine associations between clinical scores and T cell effector functions, we separated T scores from NIH Toolbox or PROMIS-57 measurements (Fig. 1C-D) into quartiles and used only the lowest and highest groups (Q1 vs. Q4) for analysis (Fig. S9A, red boxes). Neuro-PASC subjects reporting high degrees of pain produced significantly more IL-6 and less cytotoxic effector molecules from CD8^+^ T cells than those with low pain scores (Fig. 5C-D). Further, patients with low depression scores had virus-specific CD8^+^ TEM that expressed higher levels of perforin, while those reporting high scores had elevated granzyme production (Fig. 5E, F). Cognitive impairment was also significantly correlated with T cell responses.

**Figure 5:**
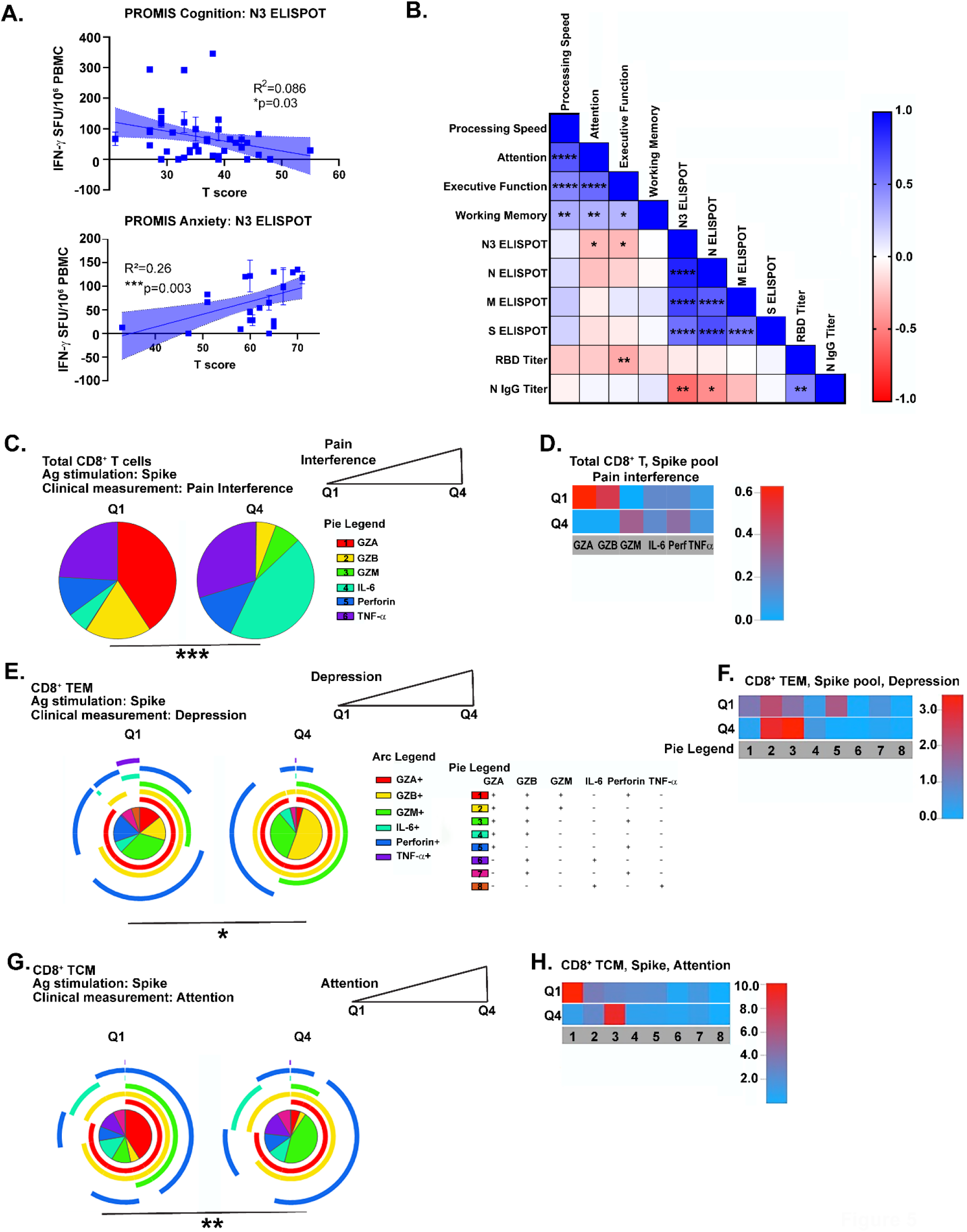
Correlation of cognitive and psychiatric clinical measures with virus-specific immune responses in Neuro-PASC patients. A.) T cell production of IFN-γ against the C-terminal region of N protein is negatively correlated with self-reported cognition scores (top) and positively correlated with anxiety scores (bottom) in Neuro-PASC patients. B.) Correlation matrix showing significant positive (blue) or negative (red) links between adaptive immune responses and clinical parameters in Neuro-PASC patients. C.) Neuro-PASC patients with high Pain Interference scores express more IL-6 from CD8^+^ T cells in response to S peptides. D.) Heatmap representing data in (C). E.) Neuro-PASC patients with high depression scores have CD8^+^ TEM that express higher levels of cytolytic effector molecules in response to S peptides. F.) Heatmap representing data in (E). G.) Spike-specific CD8^+^ TCM from Neuro-PASC patients with high executive function cognitive scores express less granzyme M compared with those with low scores. H.) Heatmap of data in G. Data representative of 5 independent experiments with n=39-51 for correlation data analysis (A-B) and n=8-9 NP subjects per quartile for SPICE analysis (C-H). Correlations calculated using simple linear regression (A) or nonparametric Spearman rank correlations (B). All pie graphs are background subtracted (unstimulated conditions). *p<0.05, **p<0.01, ****p<0.001 using Permutation tests.

Patients scoring low on attention by NIH Toolbox had CD8^+^ T central memory cells (TCM) expressing different patterns of cytolytic effectors compared to those with high attention scores (Fig. 5G-H). Similar analyses were performed for other memory T cell subsets, and significant differences were also found in correlations with processing speed, working memory, and global pain (Fig. S9B-K).

### Elevated immunoregulatory proteins and decreased antiviral and T_h_1-response proteins in Neuro-PASC patients correlate with cognitive dysfunction

The multiplexed proteomics platform SOMAscan has been successfully used in previous studies to identify biomarkers associated with conditions such as hepatocellular carcinoma (30), Alzheimer’s disease (31), and drug treatment of myocardial infarction (32). The technology utilizes the natural 3D folding of single-stranded DNA-based protein recognition aptamers to quantify levels of more than 7000 unique proteins in biological fluids (33). We used this platform to determine whether Neuro-PASC patients had proteomic signatures distinct from COVID convalescents through pathway analysis as well as comparison of individual protein levels. Gene set enrichment pathway analysis (GSEA) has previously been used on proteomics data to identify dysregulated circuits in Duchenne muscular dystrophy (34). GSEA similarly identified an enrichment in immunoregulatory pathway proteins in Neuro-PASC patients and conversely, elevated antiviral and T_h_1-type immune pathways in COVID convalescents (Fig. 6A). Comparison of individual proteins enriched in the immunoregulatory pathway identified significantly elevated CD247, SIGLEC7, MICA, and other molecules involved in regulating T cell activation (Fig. 6B, top panel). In contrast, plasma from healthy COVID convalescents were enriched in the antiviral TASOR pathway proteins H2AC11, METTL3, and MAP4K1 (Fig. 6B, bottom panel), among others, which are involved in preventing intracellular viral replication and T cell differentiation (35). A number of proteins were correlated with cognitive performance or neurologic symptom severity in both pathways (Fig. 6C), with a particularly significant negative correlation between self-reported cognition scores and expression of the inhibitory NK cell/CD8^+^ T cell receptor KLRC1 (Fig. 6C, left panel).

**Figure 6:**
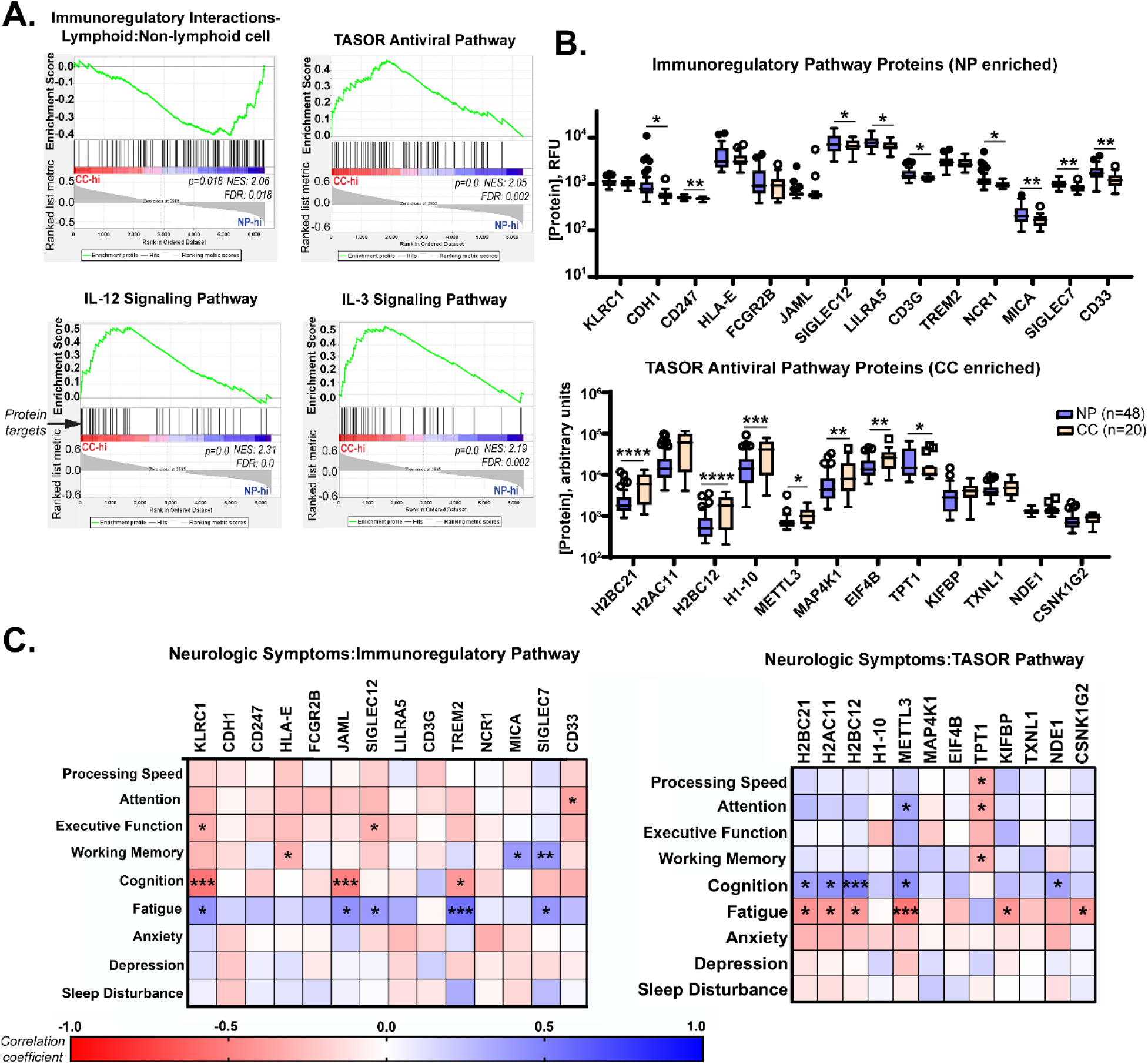
Neuro-PASC patients have elevated levels of immunoregulatory proteins in plasma that are correlated with symptom severity and cognitive dysfunction. A.) Gene set enrichment analysis (GSEA) of proteomic data demonstrating elevations in immunoregulatory pathway-related proteins (top left panel) in Neuro-PASC patients and elevated pro-inflammatory and antiviral pathway-related proteins (top right, bottom panels) in healthy COVID convalescents. List of proteins analyzed in each pathway found in Tables S6-S9. B.) Comparison of concentrations of individual immunoregulatory (top) and TASOR antiviral pathway-associated proteins (bottom) between Neuro-PASC patients and healthy COVID convalescents. C.) Patient-reported outcomes of symptom severity and cognitive metrics are significantly correlated with expression levels of immunoregulatory proteins (left) and TASOR pathway proteins (right). RFU: relative fluorescence units. FDR: false discovery rate. NES: normalized enrichment score. *p<0.05; **p<0.01; ***p<0.005; ****p<0.0001 by Student’s t test (B) or Pearson correlation (C).

Overall, our study demonstrates that Neuro-PASC patients have elevated IFN-γ responses to internal (Nucleocapsid & Membrane) proteins of SARS-CoV-2, enhanced activation of Tfh cells linked to increased anti-Nucleocapsid antibody production, but impaired activation of CD8^+^ memory T cells compared with COVID convalescents. In addition, we show unique correlations between the severity of cognitive deficits or quality of life impairments and increased virus-specific T cell responses, suggesting that higher T cell responses are not always linked to better clinical outcomes. Importantly, proteomics analysis found upregulations in immunoregulatory pathway proteins and downregulation in inflammatory and antiviral response proteins in Neuro-PASC patients that were highly correlated with neurocognitive dysfunction. Together, we show that Neuro-PASC patients exhibit distinct activation and effector signatures in multiple aspects of the T cell response.

## Discussion

COVID-19 is recognized as a multi-organ disease with long-term sequelae associated with neurological dysfunction. PASC has been reported in up to 87% of those hospitalized with SARS-CoV-2 pneumonia and in 30% of those with mild disease who do not require hospitalization (36, 37). Long-term sequelae after coronavirus infections can persist for years (9); therefore, it is important to characterize immune responses associated with PASC. Prior studies have focused on acute infection in COVID convalescents broadly as opposed to focusing on those with PASC (38, 39). We aimed to fill this knowledge gap and examine how virus-specific adaptive immunity differs in patients with Neuro-PASC vs. healthy COVID convalescents.

Clinically, Neuro-PASC resembles myalgic encephalomyelitis/chronic fatigue syndrome (ME/CFS), which is often reported as a post-viral syndrome (40). The causes of ME/CFS remain elusive, and the underlying mechanisms of Neuro-PASC are similarly unknown. One hypothesis is that Neuro-PASC symptoms may be caused by direct infection of the CNS. SARS-CoV-2 may gain entry into the CNS through the olfactory bulb and has been shown to infect neurons in vitro, which is supported by viral protein expression in neurons from post-mortem autopsies and presence of virus in the brain in mouse models (41, 42). However, other studies were unable to find evidence of SARS-CoV-2 in the CNS of patients who died with neurologic symptoms (43) or in the cerebrospinal fluid (CSF) (44), suggesting that infection of the nervous system may be transient or may not occur in all infected individuals. Importantly, a study in Neuro-PASC patients did not find any SARS-CoV-2 RNA or intrathecally-produced viral antibodies in the CSF at 90 days post-infection (45). As lumbar punctures or brain biopsies are not indicated in the majority of Neuro-PASC patients seen in our clinic, reproducing the above study results in ambulatory populations is not possible. Additional hypotheses for Neuro-PASC pathogenesis include autoimmune mechanisms which are suggested by the increased ratio of females to males affected, similar to that seen in rheumatoid arthritis or other autoimmune diseases (16, 46), or the possibility of persistent SARS-CoV-2 infection (47). It is also important to mention that the majority of Neuro-PASC patients in our study had one or more comorbidities, while comorbidity rates in COVID convalescents were lower (Figure 1B). It is thus possible that the presence of comorbidities can increase the predilection for Neuro-PASC, and this combined with the heterogeneous severity of acute COVID-19 in these patients may bias our results compared with healthy COVID convalescents. However, more than half of U.S. adults are estimated to have one or more chronic health conditions (48), which may put millions of people at risk for long COVID syndrome.

T cell responses were similar between Neuro-PASC and COVID convalescent groups in response to SARS-CoV-2 S peptides (Fig. 2A, S2A). These results are expected as T cell responses to Spike protein remain diverse after both infection (11) and vaccination (49). In contrast, T cells from Neuro-PASC patients retained high IFN-γ responses to N and M peptides, while COVID convalescents had limited reactivity to the C-terminal domain of N protein (Fig. 2C). Further characterization localized the enhanced Neuro-PASC T cell reactivity to amino acids 309-402 of N protein (Fig. 2D). It is possible that Neuro-PASC T cell responses remain high towards N and M protein due to altered T cell clonal expansion patterns compared with COVID convalescents. Studies have identified SARS-CoV-2 specific T cell receptor (TCR) sequences that are clonally expanded in a large population of COVID convalescents (50). However, these studies did not determine whether convalescent subjects had PASC at the time of sample collection. Further studies are needed to determine whether the enhanced T cell responses to N and M proteins in Neuro-PASC patients correspond to differential patterns of virus-specific TCR clonal expansion which may inform vaccination and treatment strategies.

T follicular helper (Tfh) cells can be important for the elicitation of antibody responses after SARS-CoV-2 infection by helping to establish germinal centers in secondary lymphoid organs, ultimately resulting in the production of high-affinity antibodies (51). SARS-CoV-2 N peptides activated Tfh cells in Neuro-PASC but not COVID convalescent groups (Fig. 3A-B), while the opposite pattern was observed in response to S and M peptides. Tfh responses are also dependent on antigen levels and directly correlate with the magnitude of the antibody response (52). Indeed, Neuro-PASC patients with high N-specific Tfh activation also displayed elevated antibody titers compared to controls (Fig. 3C). This was despite the fact that we obtained their samples more than 6 months after acute infection when anti-N antibody titers would fall beyond detection in most COVID convalescents (53).

Effective generation of T cell memory responses can be important to protect against future infections with the same pathogen. CD8^+^ T effector memory (TEM) cells from Neuro-PASC patients displayed reduced antigen-specific activation compared with COVID convalescents (Fig. 4A-B), suggestive of a diminished effector response. CD137 may play a role here as costimulation provides an important signal necessary for the activation of virus-specific T cells (54), but this costimulatory marker was reduced on CD8^+^ memory T cells from Neuro-PASC patients relative to COVID convalescents (Fig. 4A-4B). Prior studies have shown that asymptomatic individuals display a robust T cell response to SARS-CoV-2 Nucleocapsid protein after infection (39), suggesting that the lack of T cell memory responses in Neuro-PASC patients is detrimental.

We also observed a significant elevation in CD8^+^ TEMRA cells in Neuro-PASC patients compared to control groups (Fig. 4C). CD8^+^ TEMRA cells have been shown to accumulate during persistent viral infections and contribute to immunosenescence (55). Their decreased virus-specific activation in Neuro-PASC patients (Fig. 4E) suggests lower cytotoxic capacity compared with healthy COVID convalescents and coincides with their elevated production of granzyme in Fig. 4I. Our data suggest that CD8^+^ TEMRA cells may be functionally anergic in Neuro-PASC patients compared with COVID convalescents and may contribute to the pathogenesis of PASC.

Significantly, CD8^+^ TEM from Neuro-PASC patients expressed higher levels of IL-6 in response to viral peptides, while in contrast CD4^+^ T cells from COVID convalescents expressed higher IL-6 (Fig. 4F-G, 3E). Clinically, CD8^+^ T cell production of IL-6 was also significantly correlated patient-reported pain scores (Fig. 5C). IL-6 plays opposing regulatory roles in T cell memory responses during viral infections. CD4^+^ memory T cells require IL-6 for activation and proliferation during viral infection (56). Thus, it is possible that elevated IL-6 production by CD4^+^ T cells in COVID convalescents is a correlate of protective anti-viral immunity. However, IL-6 can also suppress T_H_1 differentiation (57) and was found to promote pathogen survival while exacerbating clinical disease during SARS-CoV-1 infection (58). In fact, blocking IL-6 activity enhances virus-specific CD8^+^ T cell immunity (59), and overexpression of IL-6 can lead to viral persistence by impairing CD8^+^ lytic functions (60) and the development of CD8^+^ T cell memory (61). Indeed, severely ill COVID-19 patients had high serum levels of IL-6 that significantly correlated with poor clinical outcomes (38). Thus, our data suggest that enhanced IL-6 production by CD8^+^ T cells and/or innate immune cells (Fig. S8A-B) may be involved in the etiology or pathogenesis of Neuro-PASC and reveal new avenues of research for the treatment of long COVID through limiting IL-6 activity.

Neuro-PASC patients reported significantly elevated levels of anxiety, depression, pain, and other symptoms compared with COVID convalescents (Fig. 1C). The severity of these deficits was correlated with Ag-specific enhancements in polyfunctionality but decreases in polarization of memory T cell subsets (Fig. 5, S9). It is possible that T cell activation contributes to some of these symptoms. Studies in rodents have shown that T cell activation can affect the severity of pain and analgesia (62); it may follow that aberrant T cell activation can be linked to high pain scores. Inflammation-related transcriptional programs are also differentially regulated in T cells from patients with depression (63), providing a possible link between enhanced granzyme production and elevated depression scores (Fig. 5E, S9B). T cell-derived cytokines can also impact learning and memory. Mouse models of West Nile and Zika viral encephalitis have demonstrated that IFN-γ production from CD8^+^ T cells in the brain is responsible for neuronal apoptosis and spatial learning deficits (64). The association of SARS-CoV-2-specific cytokine signatures with the severity of cognitive and quality of life deficits in Neuro-PASC patients may therefore provide some predictive value in terms of clinical outcomes.

Proteomic analysis of patient plasma demonstrated that Neuro-PASC patients had relatively blunted levels of pro-inflammatory and antiviral response-associated proteins compared to COVID convalescents, while simultaneously having elevated immunoregulatory protein expression (Fig. 6A). Further analyses at the individual protein level showed upregulation of immunoregulatory proteins such as CD33 and NCR1 in Neuro-PASC plasma, involved in T cell immunosuppression in acute myeloid leukemia (65) and suppression of antiviral CD8^+^ T cell responses (66), respectively. These data support our findings showing decreased antiviral CD8^+^ TEM and TEMRA responses (Fig. 4) and suggest that an imbalance between immunoregulatory and antiviral pathways may play a role in Neuro-PASC pathogenesis. In line with this, one of the strongest associations we found with poor cognitive scores involved the NK and CD8^+^ T cell inhibitory receptor KLRC1 that downregulates cytotoxic capacity (67) (Fig. 6C). KLRC1 expression on CD8^+^ T cells is upregulated by IL-6 (68), and enhanced KLRC1 expression has been found on exhausted CD8^+^ T cells from acute COVID-19 patients (69). Based on our data, it is therefore possible that enhanced IL-6 production from CD8^+^ T cells in Neuro-PASC patients may upregulate KLRC1 and suppress CD8^+^ T cell function, which may impact Neuro-PASC symptom severity. Together, these data illuminate a specific T cell signature associated with of Neuro-PASC.

## Limitations of study

One limitation is the relatively small sample size of unvaccinated COVID convalescent subjects. This was due to the wide implementation of SARS-CoV-2 vaccines in Chicago area soon after beginning study enrollment. Another limitation was not being able to control for time of sample collection with respect to date of COVID-19 symptom onset. Additionally, as we hypothesize that Neuro-PASC could be the result of a persistent or protracted infection, future studies would require testing of potential cryptic reservoirs, including stool or CNS tissue from Neuro-PASC patients.

## Methods

### Study design

We attempted to include a robust sample size for every patient group, including those in the vaccine portion of the study. Enrollment sizes for the COVID convalescent group in particular was limited despite posting recruitment flyers as well as on social media due to the widespread rollout of vaccines. Data collection was stopped in our study at the indicated endpoints. Data inclusion/exclusion criteria are described below in the *Study participants* section. Endpoints were selected prospectively. Replicates for each experiment are described in figure legends.

Research objectives were to identify and characterize T cell responses to SARS-CoV-2 linked to Neuro-PASC pathogenesis and specify how these responses differed from COVID convalescents without lasting symptoms. We enrolled Neuro-PASC outpatients, COVID convalescents, and unexposed healthy controls for our study. Experimental design is outlined in Fig. 1A. Subjects were not randomized and investigators were not blinded to the study subjects’ grouping prior to conducting experiments and analyzing data.

### Study participants, NIH Toolbox, and PROMIS-57 data collection

We enrolled consenting adult outpatients seen in the Neuro-PASC-19 clinic at Northwestern Memorial Hospital from September 2020-September 2021, including 89 Neuro-PASC patients with documented PCR+ or seropositive IgG results for SARS-CoV-2. In parallel, we recruited 43 COVID convalescents from the surrounding community who tested either PCR+ or seropositive for SARS-CoV-2 before vaccination but had no lingering neurological symptoms and 34 healthy controls who tested PCR- for SARS-CoV-2 and were also seronegative for IgG against SARS-CoV-2 Spike RBD prior to vaccination. All study subjects remained living throughout the period of observation. Heparinized blood samples were collected one time from each subject at an average of 161.5-223.0 days post-symptom onset (as in Fig. 1B). Other demographic information, including comorbidity information, is contained in Fig. 1B and Supplementary Tables 2-6. Neuro-PASC patients completed a cognitive function evaluation in the clinic coincident or near the date of their blood sample acquisition with the National Institutes of Health (NIH) Toolbox v2.1 instrument, including assessments of: processing speed (pattern comparison processing speed test); attention and executive memory (inhibitory control and attention test); executive function (dimensional change card sort test); and working memory (list sorting working memory test) (19). PROMIS-57 patient-reported quality of life assessments were administered to Neuro-PASC and COVID convalescent subjects an average of 72 days post-sample collection. Both PROMIS-57 and NIH Toolbox results are expressed as T-scores with a score of 50 representing the normative mean/median of the US reference population and a standard deviation of 10. Toolbox results are adjusted for age, education, gender, and race/ethnicity. Lower cognition T-scores indicate worse performance while higher fatigue, depression, anxiety, or pain interference T-scores indicate greater symptom severity.

### PBMC and plasma collection

30mL of venous blood from study volunteers was collected in blood collection tubes containing sodium heparin from BD Biosciences. Whole blood was layered on top of 15mL of Histopaque 1077 (Sigma-Aldrich) in 50mL Leucosep blood separation tubes (Greiner Bio-One) and spun at 1000g for 18min at RT. Plasma was collected and stored at −80°C. The PBMC layer was collected and washed 2x in sterile PBS before red blood cell lysis with ACK buffer (Quality Biologicals). PBMCs were used in assays either immediately or frozen down for use in the near term.

### SARS-CoV-2 peptide antigens

All S, N and M peptide arrays used in ELISPOT and flow cytometry studies were obtained from BEI Resources, NIAID, NIH: Peptide Array, SARS-Related Coronavirus 2 Spike (S) Protein; NR-52402, Nucleocapsid (N) Protein, NR-52404; Membrane (M) Protein, NR-52403. The S peptide array consisted of 181 peptides of 13-17aa in length and split into 6 sub-pools (S1-S6) containing 30-31 peptides each. The N peptide array consisted of 59 peptides of 13-17aa each split into 3 sub-pools containing 29-30 peptides each (Fig. 2B) or with 1 sub-pool further divided into 5 pools of 3-4 peptides each (Fig. 2D). The M peptide array consisted of 31 peptides of 12-17aa; details in Fig. S1. All peptides were dissolved in either sterile H_2_O or 50% sterile H_2_O-DMSO up to 1mL for a universal 1mg/mL stock concentration. Peptides were used at a final concentration at 2μg/mL in all assays.

### IgG Spike RBD and Nucleocapsid ELISA

Antigen-specific total antibody titers were measured by ELISA as described previously(70). In brief, 96-well flat-bottom MaxiSorp plates (Thermo Scientific) were coated with 1 µg/ml of Spike RBD for 48 hr at 4°C. Plates were washed three times with wash buffer (PBS + 0.05% Tween 20). Blocking was performed with blocking solution (PBS + 0.05% Tween 20 + 2% bovine serum albumin), for 4 hr at room temperature. 6 µl of sera was added to 144 µl of blocking solution in the first column of the plate, 1:3 serial dilutions were performed until row 12 for each sample, and plates were incubated for 60 min at room temperature. Plates were washed three times with wash buffer followed by addition of secondary antibody conjugated to horseradish peroxidase, goat anti-human IgG (H + L) (Jackson ImmunoResearch) diluted in blocking solution (1:1000) and 100 µl/well was added and incubated for 60 min at room temperature. After washing plates three times with wash buffer, 100 µl/well of Sure Blue substrate (SeraCare) was added for 1 min. Reaction was stopped using 100 µl/well of KPL TMB Stop Solution (SeraCare). Absorbance was measured at 450 nm using a Spectramax Plus 384 (Molecular Devices). SARS-CoV-2 RBD and N proteins used for ELISA were produced at the Northwestern Recombinant Protein Production Core by Dr. Sergii Pshenychnyi using plasmids that were produced under HHSN272201400008C and obtained from BEI Resources, NIAID, NIH: Vector pCAGGS containing the SARS-related coronavirus 2, Wuhan-Hu-1 spike glycoprotein gene (soluble, stabilized), NR-52394 and receptor binding domain (RBD), NR-52309, nucleocapsid gene NR-53507.

### Cell stimulation and IFN-γ/IL-2 ELISPOT

Multiscreen-IP plates (Millipore-Sigma) were coated overnight at 4°C with 2μg/mL anti-IFN-γ (clone 1-D1K, Mabtech) or 5μg/mL anti-IL-2 (clone MT2A91/2C95, Mabtech), washed with sterile PBS, and blocked with complete RPMI-10% FBS. PBMC isolated from Neuro-PASC, COVID convalescent, and healthy control subjects were used either freshly isolated or after thawing and resting overnight in media containing 10ng/μL recombinant human IL-15 (Peprotech) at 37°C, 5% CO_2_. Cells were then plated at a concentration of 2.5×10^5^ cells/well in 100μL of media and stimulated with the indicated antigen mixtures from SARS-CoV-2 at a concentration of 2μg/mL in complete RPMI medium containing 5% human AB serum (Sigma-Aldrich) and 5ng/mL IL-15. Plates were incubated at 37°C, 5% CO_2_ for 20h and washed 5x with dH_2_O and PBS-0.05% Tween-20 (PBS-T). 2μg/mL biotinylated IFN-γ (clone 7-B6-1, Mabtech) or 5μg/mL IL-2 (clone MT8G10, Mabtech) diluted in PBS-10% FBS (PBS-F) was added to the respective wells and plates were incubated for 1.5h at RT. Plates were subsequently incubated for 40 minutes at RT in streptavidin-alkaline phosphatase in PBS-F (Jackson ImmunoResearch) was added after washing plates 5x in PBS-T. ELISPOT plates were developed using an Alkaline Phosphatase Conjugate Substrate Kit according to manufacturer’s instructions (Bio-Rad Laboratories, Carlsbad, CA). IFN-γ or IL-2-producing cells were quantified using an ImmunoSpot reader (Cellular Technologies, Ltd., Shaker Heights, OH).

### T cell receptor variable beta chain sequencing

Immunosequencing of the CDR3, V, and J regions of human TCRβ chains was performed using the immunoSEQ® and T-MAP COVID Assays (Adaptive Biotechnologies, Seattle, WA). Genomic DNA extracted from individual subject’s PBMC was amplified in a bias-controlled multiplex PCR, followed by high-throughput sequencing. Sequences were then filtered to identify and quantitate the absolute abundance of each unique TCRβ template for further analysis as previously described (49). TCR specificities to SARS-CoV-2 Nucleocapsid were determined using immuneCODE, a publicly available database accessed via the immunoSEQ Analyzer platform. Peptide antigens specific for each TCR were then aligned to the Nucleocapsid amino acid sequence to demarcate regional specificity (“N1” vs. “N2” vs. “N3”). The value for the top expanded N3-specific TCR clone was counted for each NP and CC subject and analyzed in Fig. 2E.

### Antibodies and Flow Cytometry

Fresh or frozen PBMCs isolated from the indicated patient groups were stimulated with antigen mixtures as above for 20-22h at 37°C, 5% CO_2._ For intracellular staining and cytokine detection, the Brefeldin-A Golgi plug (Biolegend) was added at a 1:1000 concentration 2 hours after antigenic stimulation commenced. Cells were washed with PBS-1% BSA after incubation and incubated with the indicated antibodies for surface phenotyping by AIM assay or for intracellular cytokine staining (ICS; antibodies used described in Supplemental Table 1). Cells from each subject were left unstimulated in medium containing 5ng/mL IL-15 (“background”) or stimulated in the presence of the indicated antigens. Fixation and permeabilization was performed using Cytofix/Cytoperm (BD Biosciences). Surface staining was done in the dark at 4°C for 30 minutes, while ICS was done in the dark at RT for 45 minutes. Flow cytometry was conducted on 2-5×10^5^ cells per condition. Data was acquired on a BD FACSymphony Spectral analyzer and analyzed using FlowJo v10 (BD Biosciences) and SPICE-Pestle(71).

### SOMAscan Profiling

Heparinized plasma from 48 Neuro-PASC patients and 20 healthy COVID convalescents whose T cell and antibody responses were characterized in this study were assayed for the presence of more than 7,000 proteins using the SOMAscan proteomics platform. The SOMAscan assay is a sensitive, high-throughput technique that uses chemically modified DNA aptamers to specifically bind and quantify proteins of interest from very small quantities of plasma (33). The assay measures a wide range of receptors, intracellular signaling proteins, growth factors, and secreted proteins. All plasma samples were analyzed at SomaLogic Operating Co, Inc. (Boulder, CO).

### SomaSCAN proteomics statistical analysis

For statistical comparison, all relative fluorescence unit (RFU) values for individual proteins were first analyzed by Gene Set Enrichment Analysis (GSEA version 4.2.3; Broad Institute; Molecular Signatures Database: hallmark, curated, KEGG, and reactome gene sets) to determine significantly enriched pathways between NP and CC groups (Fig. 6A). The false discovery rate cutoff was 0.05. RFUs for proteins belonging to a particular pathway (immunoregulatory or TASOR antiviral) that were enriched in NP or CC were then analyzed using two-tailed t-Test (Fig. 6B). Within-group correlations for Neuro-PASC symptoms with individual protein concentrations were determined using Spearman’s correlation (Fig. 6C).

### Quantification and Statistical Analysis

Statistical tests to determine significance are described in figure legends and conducted largely in Prism (GraphPad). SPICE is a data-mining software application that analyzes large FLOWJO data sets from polychromatic flow cytometry and organizes the normalized data graphically. SPICE defines a statistic for the nonparametric comparison of complex distributions based on multi-component measurements (71). For pie graphs or heatmaps generated using SPICE software analysis, statistics were determined by Permutation test following unstimulated background subtraction, with additional thresholding of 0.03% to account for noise, using SPICE-Pestle. *P*-values lower than 0.05 were considered statistically significant. Quartile stratification was performed within group for the Neuro-PASC cohort (Fig. S9A). Clinical data were collected and managed using REDCap electronic data capture tools hosted at Northwestern University Feinberg School of Medicine. All error bars on figures represent values ± SEM.

### Study approval

This study was approved by the Northwestern University Institutional Review Board (Koralnik Lab, IRB STU00212583). Informed consent was obtained from all enrolled participants. Samples were de-identified before banking.

## Data Availability

The data that support the findings of this study are available from the corresponding author, [LV], upon reasonable request.

## Data Availability

The full datasets generated in the current study are available from the corresponding author upon requests.

## Acknowledgements

We would like to thank Adaptive Biotechnologies for providing sequencing services and bioinformatics support, as well as the Flow Cytometry Core Facility at the Robert H. Lurie Comprehensive Cancer Center at Northwestern University supported by Cancer Center Support Grant (NCI CA060553) for their assistance in optimizing antibody panels and help with flow cytometry instrumentation. L.V. was supported by a T32 grant (NIAMS, T32AR007611) from the Department of Rheumatology, Northwestern University Feinberg School of Medicine. P.P.M. is supported by grants from the National Institute on Drug Abuse (NIDA, DP2DA051912) and from the National Institute of Biomedical Imaging and Bioengineering (NIBIB, U54EB027049).

## Author Contributions

Conceptualization L.V. P.P.M. and I.K; Investigation L.V., B.H., Z.O., P.H.L, N.P. and G.T.; Formal Analysis L.V., B.H., M.J., E.M.L., P.P.M. and N.P.; Resources L.V., G.T., P.P.M., I.K., Data Curation L.V., E.G., J.R.C.; Writing L.V. with feedback from all authors; Supervision P.P.M and I.K.; Project Administration L.V.; Funding Acquisition L.V., P.P.M, and I.K.

## Materials and Correspondence

Please address all inquiries to Dr. Igor Koralnik or Dr. Lavanya Visvabharathy.

