## Supplemental Figures & Tables for "T cell responses to SARS-CoV-2 in people with and without neurologic symptoms of long COVID"

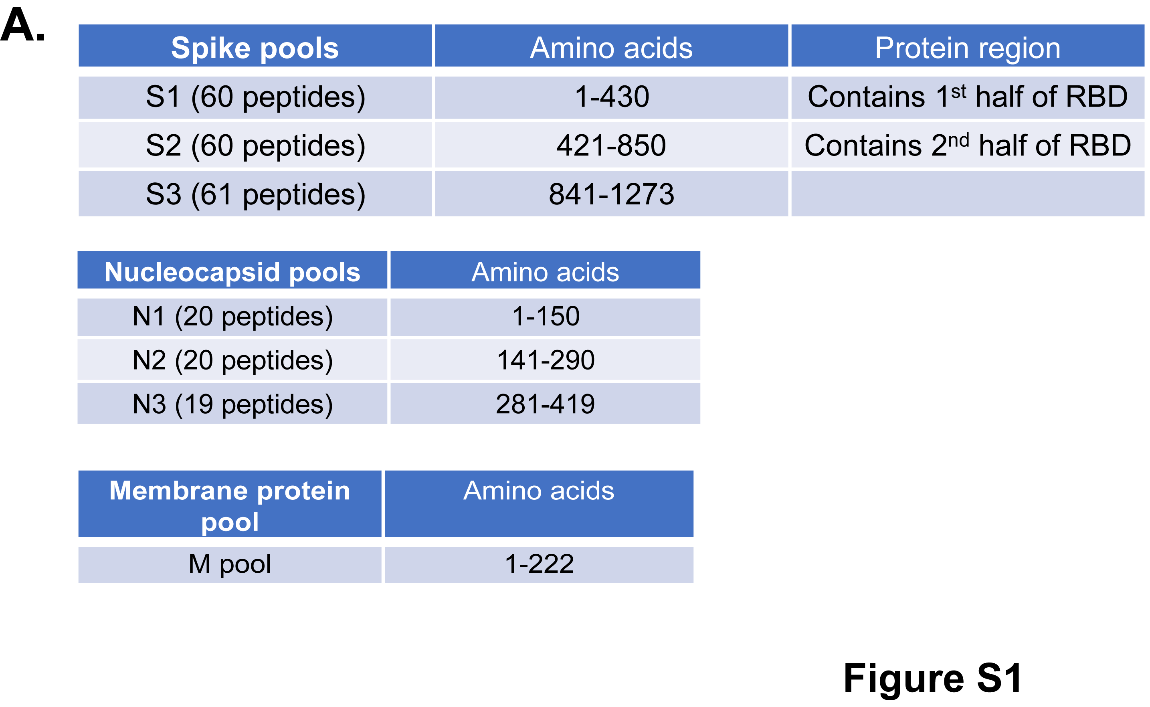

**Figure S1: SARS-CoV-2 peptides used in study**

Peptide pools derived from S, N, and M proteins from USA-WA1/2020 strain of SARS-CoV-2. S1 and S2 peptide pools from Spike protein contain the first and second halves of the Spike receptor-binding domain (RBD), respectively.

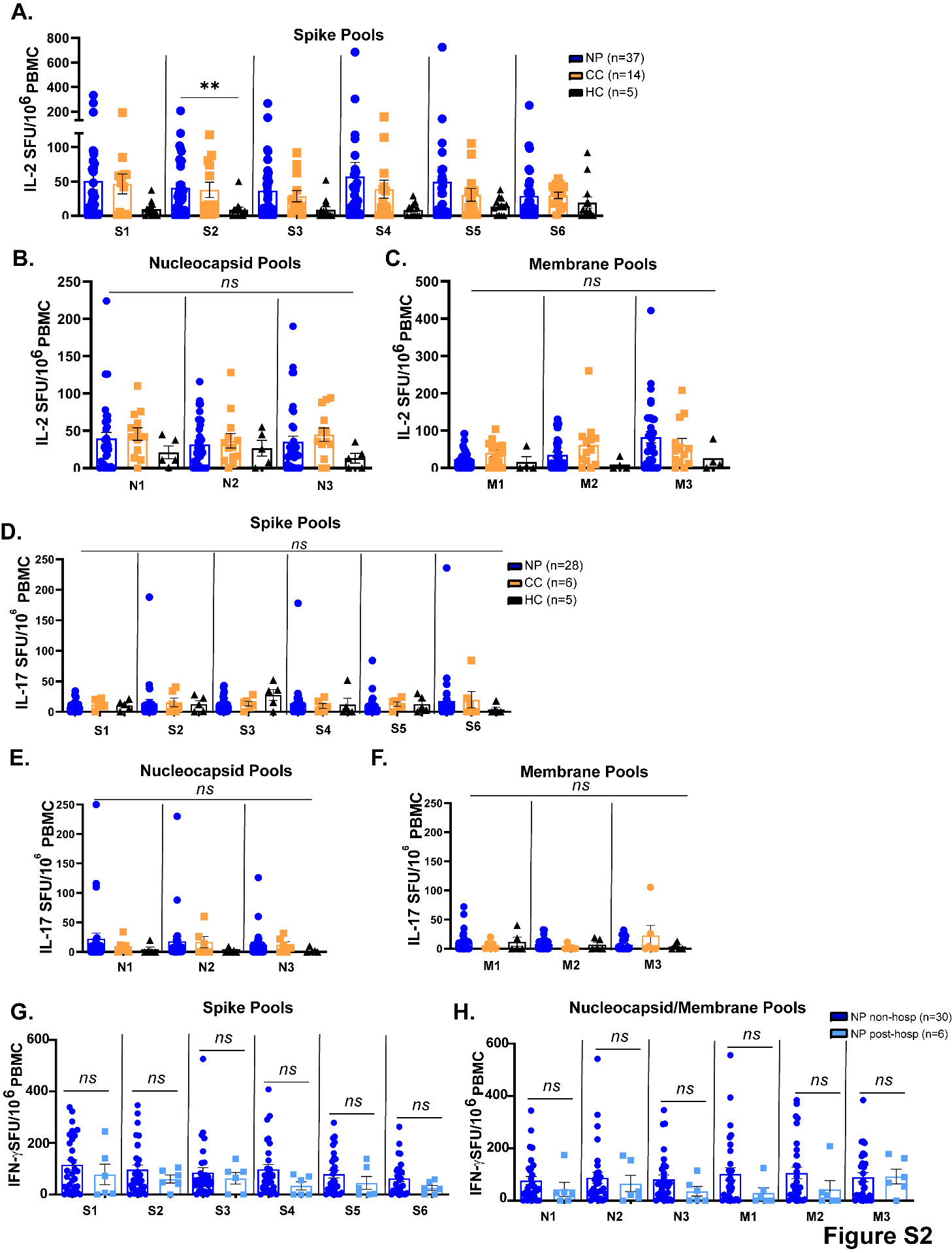

**Figure S2: IFN-γ response to Spike protein in non-hospitalized and post-hospitalized Neuro-PASC patients; IL-2 and IL-17 ELISPOT**

A.) NP and CC groups display similar IL-2 responses to peptides from SARS-CoV-2 Spike protein by ELISPOT. B-C.) N- and M- specific IL-2 production did not significantly differ between subject groups. D-F). IL-17 production in response to Spike (D), Nucleocapsid (E) and Membrane peptides (F) did not differ between groups. G-H.) Non-hospitalized and post-hospitalized Neuro-PASC patients did not show differences in the magnitude of IFN-γ production by ELISPOT after stimulation with SARS-CoV-2 Spike pools (G) or Nucleocapsid and Membrane pools (H). Data representative of 10 experiments with all conditions plated in duplicate and the indicated n values. *p<0.05, **p<0.01, ***p<0.005, ****p<0.0001 by two-way ANOVA with Tukey’s posttest.

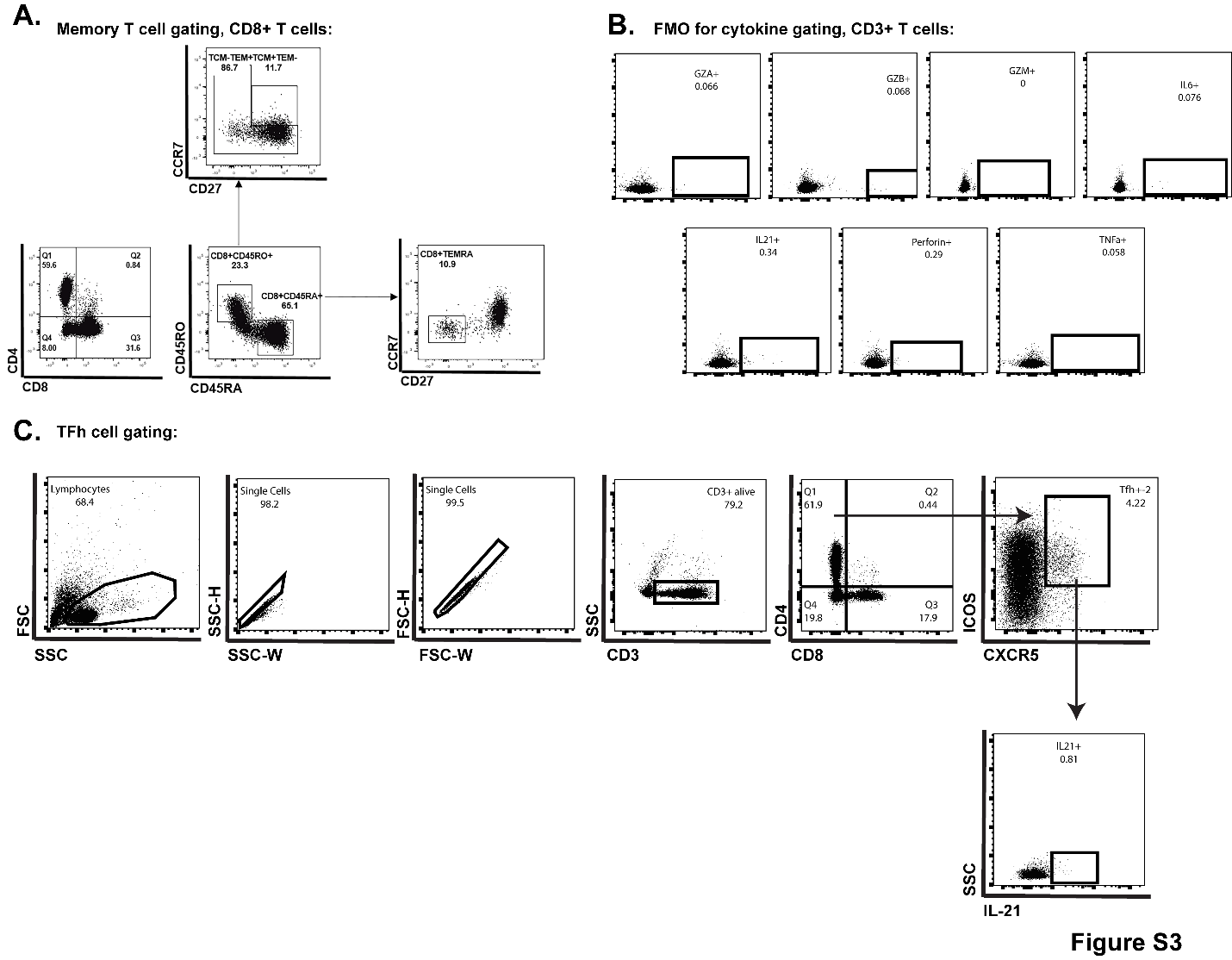

**Figure S3: Gating strategies and fluorescence-minus-one (FMOs) for cytokine production**

A.) Gating strategy for memory T cells by CD45RA/RO, CCR7, and CD27 expression. B.) FMOs used for gating in determining cytokine positive T cells. C.) Gating strategy for CD4^+^ Tfh cells and IL-21 production.

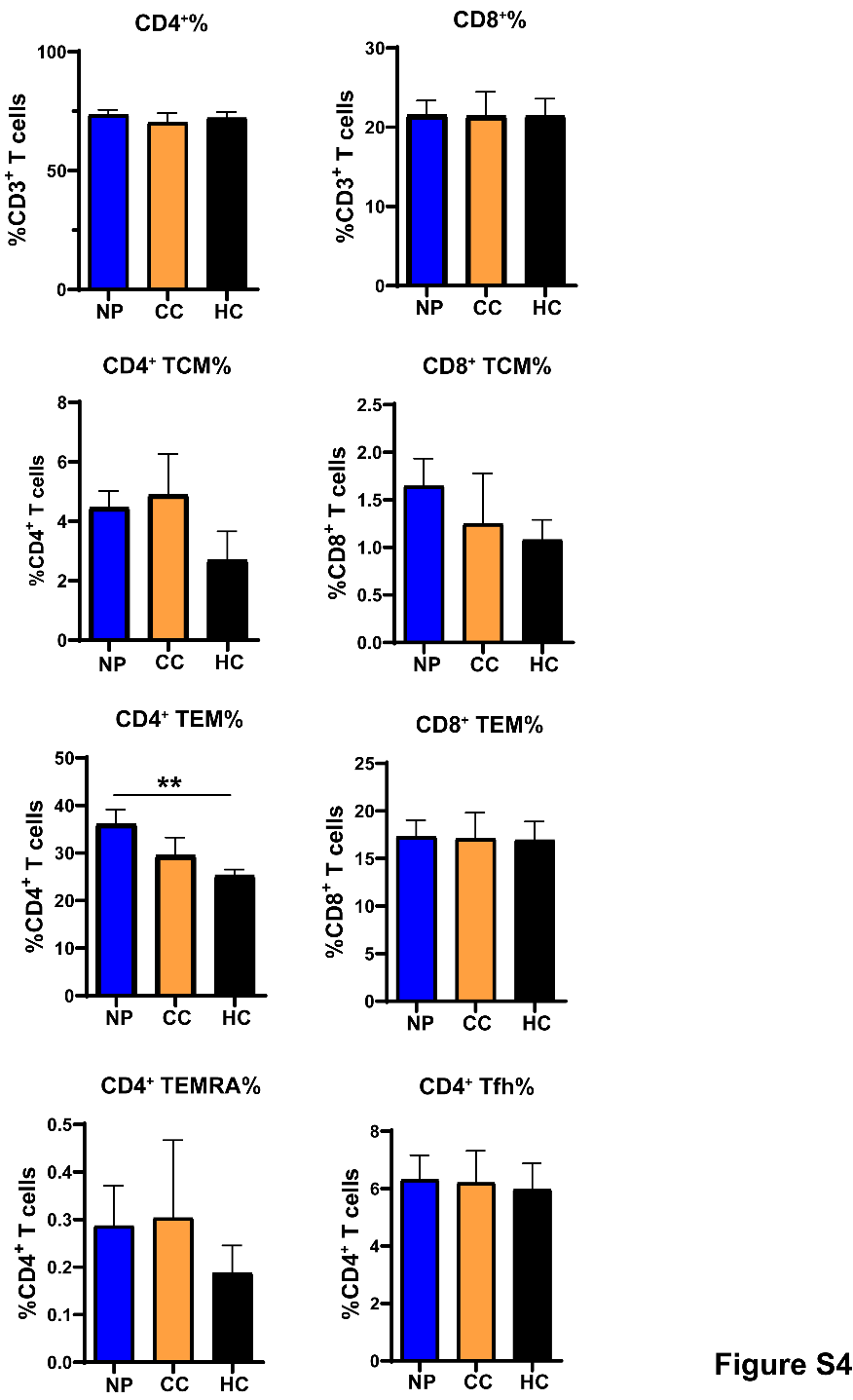

**Figure S4: Total percentages of unstimulated CD4^+^ and CD8^+^ T cell subsets between groups**

NP, CC, and HC groups displayed no significant differences in percentages of CD8^+^ TCM or TEM cells. While there was a significant increase in CD4^+^ TEM cells as a percentage of total CD4^+^ T cells in NP vs. other groups, no other cell subset including CD4^+^ TCM, TEMRA, or Tfh cells were significantly different between groups.

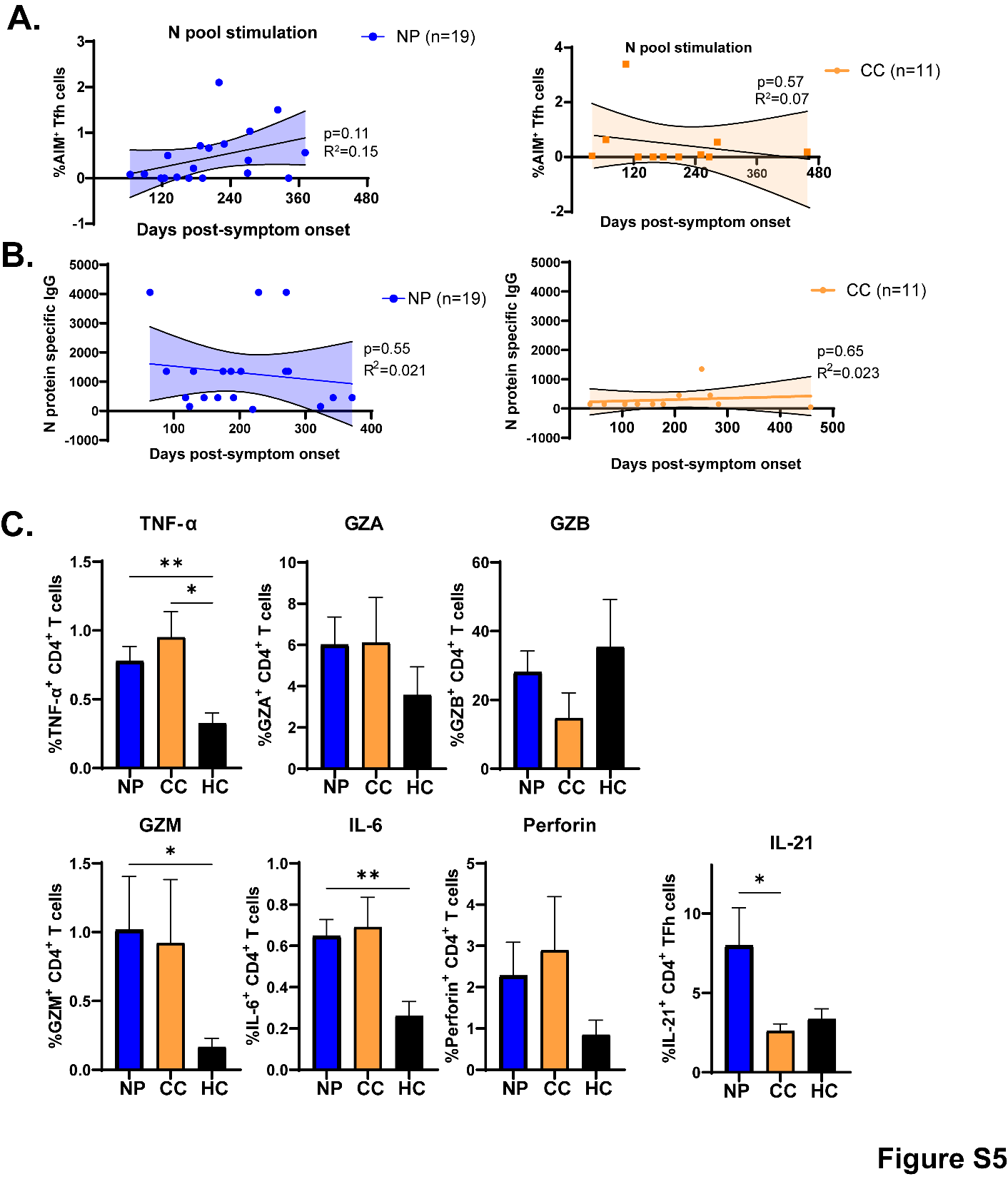

**Figure S5: Correlations between N-specific Tfh cell activation or IgG titers and days post-infection; cytokine production in unstimulated T cells**

A.) No correlation was observed between the magnitude of N-specific Tfh cell activation and the time post-symptom onset. B.) Regression analysis of N-specific IgG titers compared to time post-symptom onset showing no significant correlations. C.) Magnitude of various T_H_1-type cytokines and cytotoxic granules produced as a percentage of total CD4^+^ T cells in the unstimulated condition between NP, CC, and HC groups. IL-21 production in unstimulated CD4^+^ Tfh cells is significantly elevated in NP over CC and HC subjects. Data representative of 10 experiments with a total of n=19 NP, 11 CC, and 9 HC. *p<0.05, **p<0.01 by two-tailed Student’s t test with Welch’s correction.

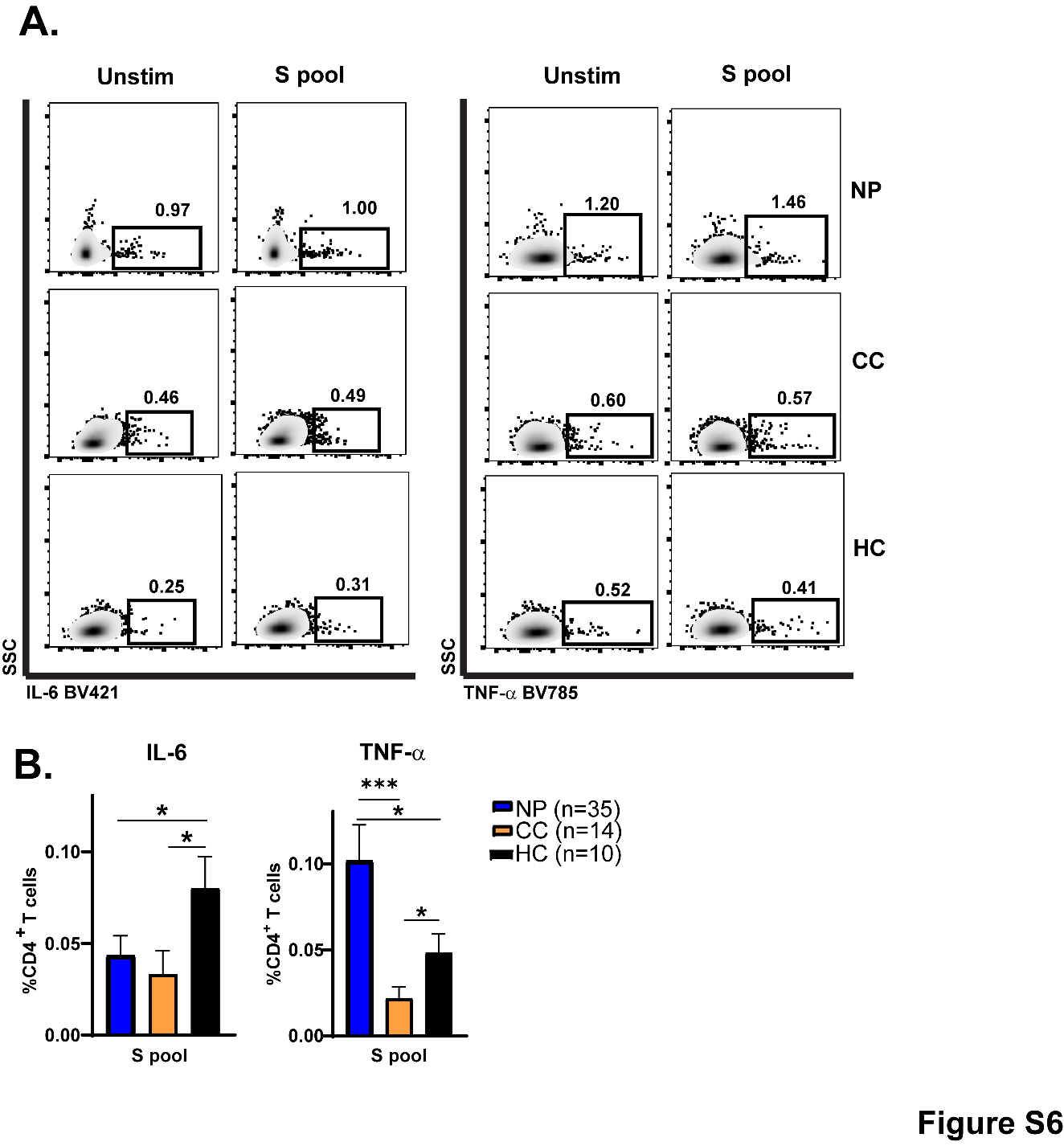

**Figure S6: IL-6/TNF-α production from CD4+ T cells after Spike peptide stimulation**

A.) Representative FACS plots showing IL-6 (left panels) and TNF-α (right panels) production from total CD4^+^ T cells in NP, CC, and HC subjects. B.) Quantification of data in A. Data combined from 6 independent experiments with the indicated n values. *p<0.05, ***p<0.005 using two-tailed Student’s t test with Welch’s correction.

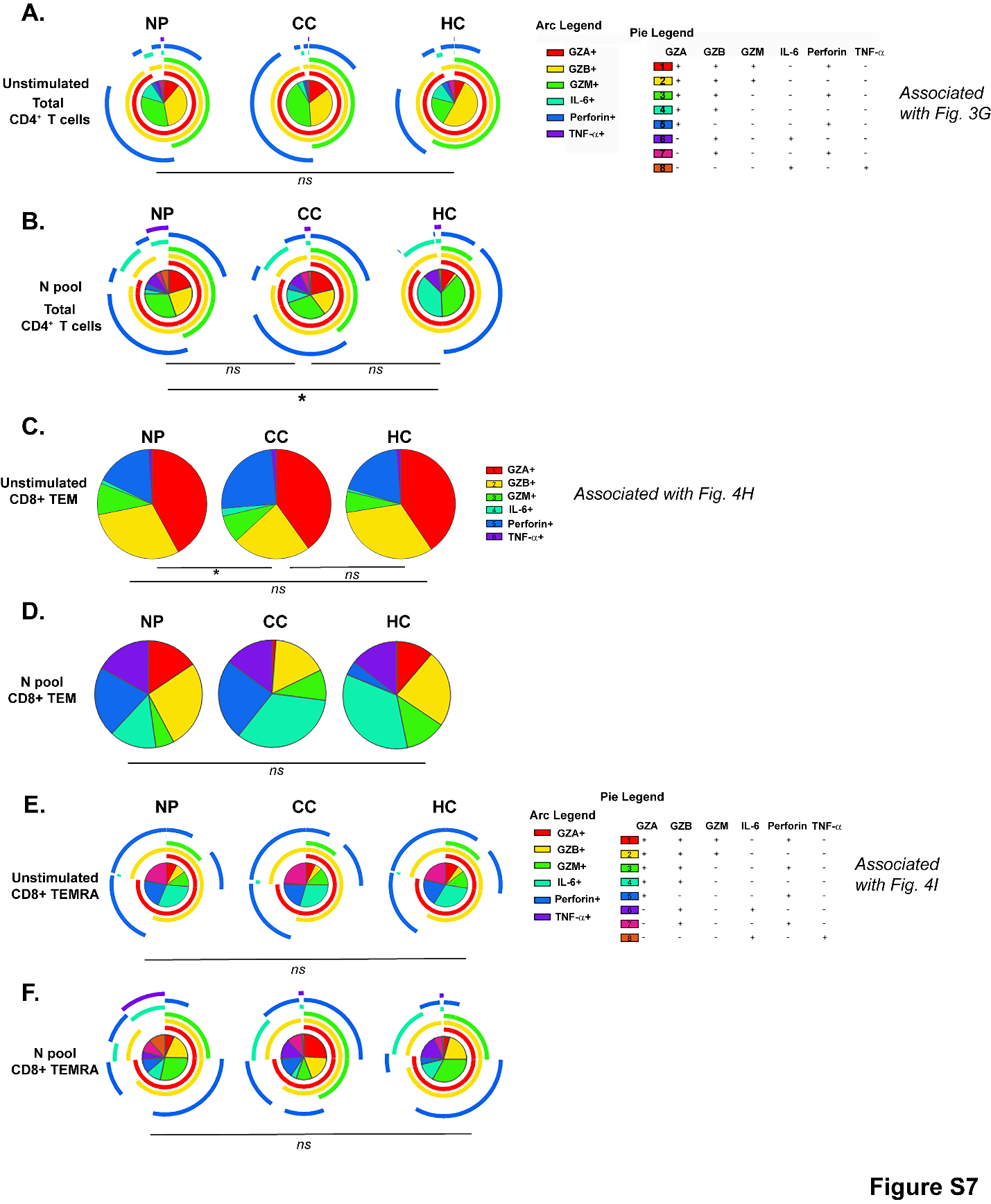

**Figure S7: SPICE analysis on unstimulated Nucleocapsid peptide-stimulated T cell subsets**

A.) No differences in unstimulated CD4^+^ T cell polyfunctionality exist between NP patients, CC, and HC subjects (associated with Fig. 3G). B.) CD4^+^ T cell polyfunctionality after N peptide stimulation in NP patients, CC, and HC subjects. C) Unstimulated CD8^+^ TEM from NP patients produce slightly more granzyme B than CC subjects (associated with Fig. 4H). D.) No significant differences were observed in cytokine production from CD8^+^ TEM after N peptide stimulation between groups. E.) Unstimulated CD8^+^ TEMRA cells have no differences in polyfunctionality between NP patients and CC subjects (associated with Fig. 4I). Data combined from 5 independent experiments with n=34 NP, n=11 CC, HC n=9. *p<0.05 by Permutation test.

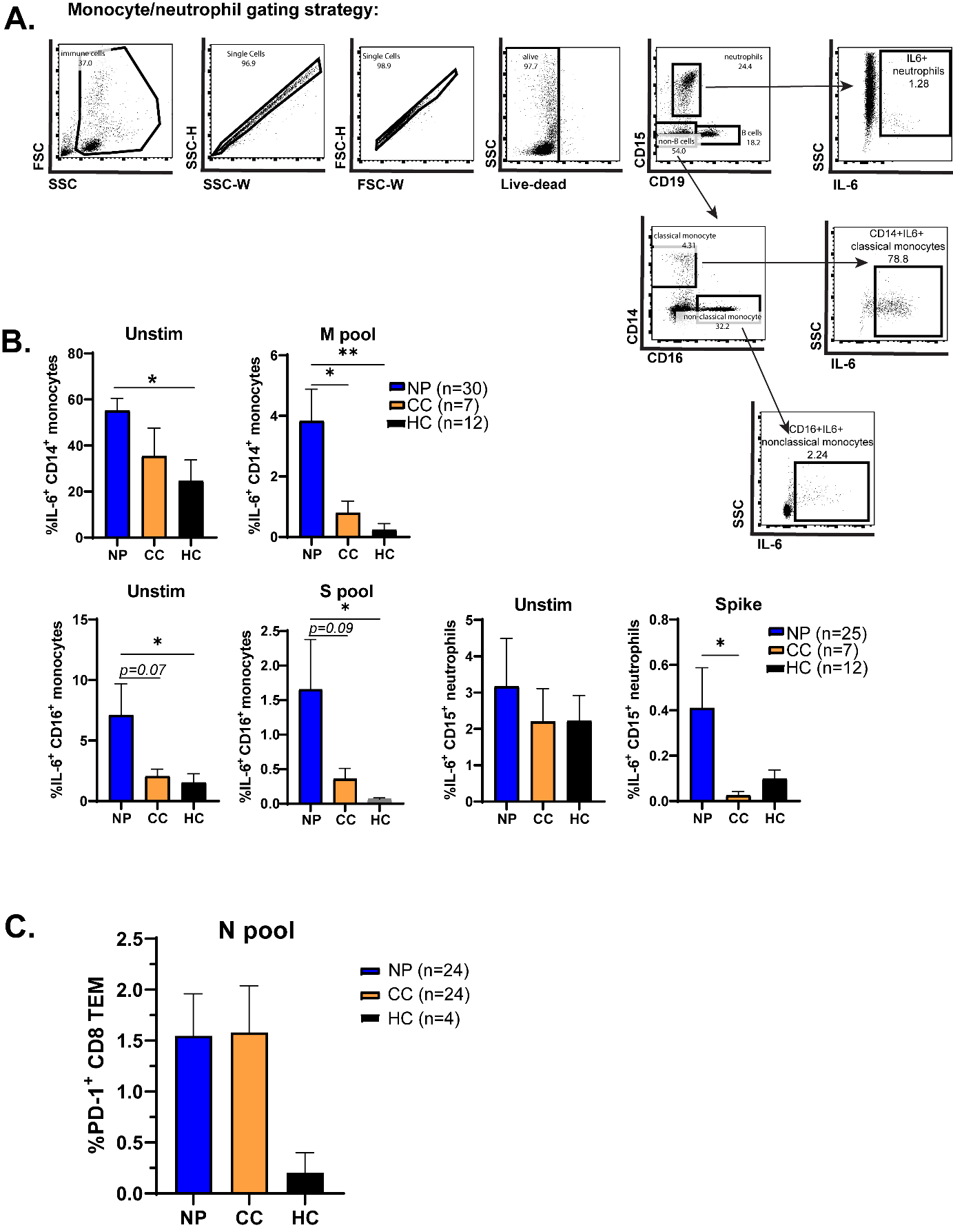

**Figure S8**

**Figure S8: Monocyte/neutrophil production of IL-6; CD8^+^ T cell exhaustion**

A.) Gating strategy for quantifying IL-6 expression in classical CD14^+^ and nonclassical CD16^+^ monocytes and in CD15^+^ neutrophils. B.) Elevated IL-6 production in S- and M-peptide stimulated monocytes and neutrophils from Neuro-PASC patients. C.) Expression of the exhaustion marker PD-1 on CD8^+^ TEM cells did not differ between Neuro-PASC patients and COVID convalescents after SARS-CoV-2 peptide stimulation. Data representative of 5 individual experiments with the indicated n. *p<0.05, **p<0.01 by two-tailed Student’s t test with Welch’s correction.

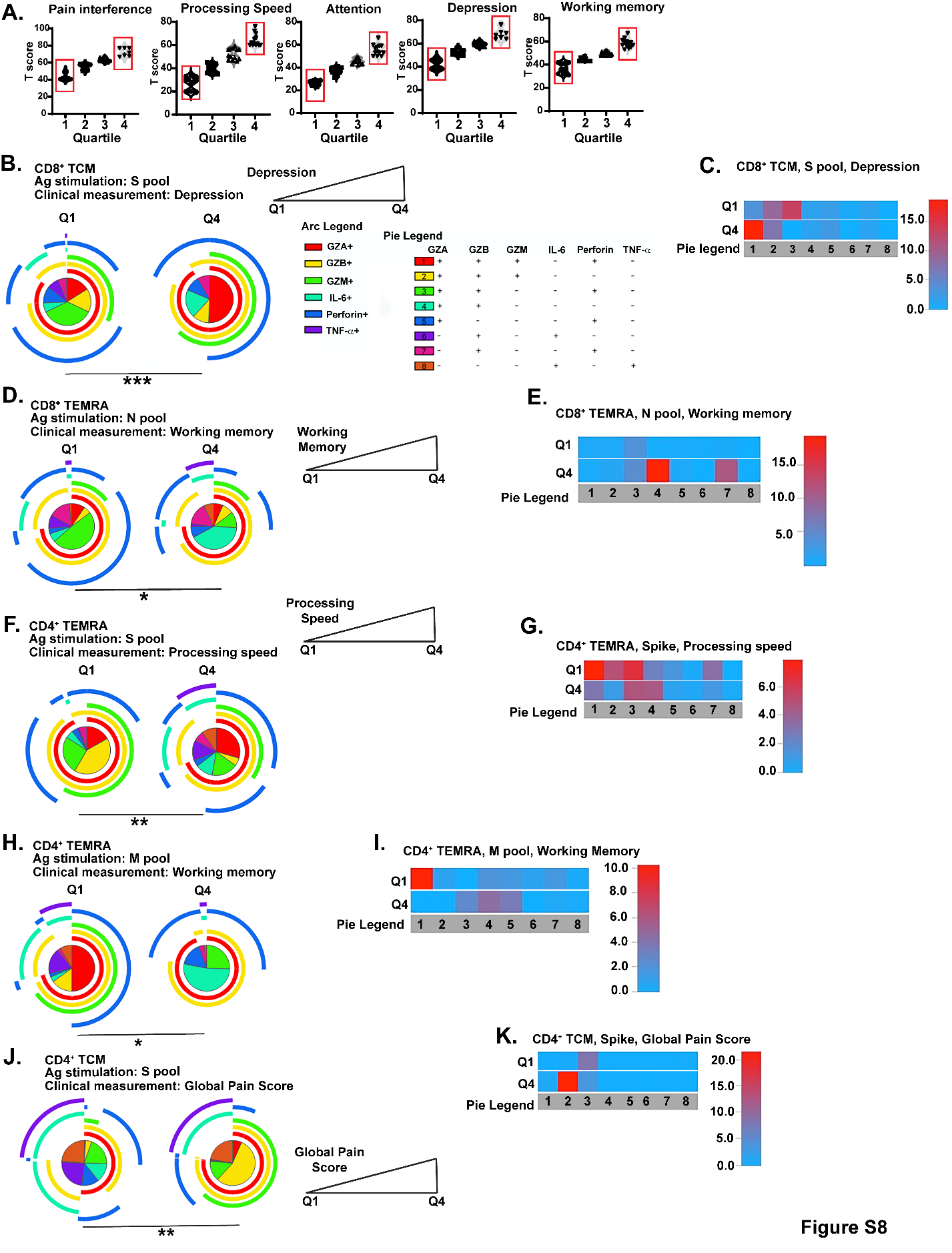

**Figure S9**

**Figure S9: Neuro-PASC NIH Toolbox and PROMIS-57 score correlations with T cell responses to SARS-CoV-2**

A.) Separation of Neuro-PASC patients into quartiles based on T scores for individual parameters from NIH Toolbox or PROMIS-57 patient-reported surveys. Only quartiles 1 and 4 (red boxes) were used as comparison groups for SPICE analyses in B-K and Fig. 5. B.) Neuro-PASC patients reporting high depression scores have significantly enhanced production of granzymes and Perforin from CD8^+^ TCM compared with those with low depression scores after Spike peptide stimulation C.) Heatmap of data in B. D.) Neuro-PASC patients with high cognitive scores for working memory had enhancement of N protein-specific polyfunctional CD8^+^ TEMRA cells producing granzymes A/B. E.) Heatmap of data in E. F.) Neuro-PASC patients scoring low on processing speed tests had more polyfunctional CD4^+^ TEMRA responses to Spike peptides than those scoring high. G.) Heatmap of data in F. H.) CD4^+^ TEMRA cells produce more granzymes A/B after M pool stimulation in Neuro-PASC patients who scored higher on the working memory test. I.) Heatmap of data in H. J.) Neuro-PASC patients reporting high levels of pain have polyfunctional CD4^+^ TCM significantly polarized towards granzyme A/B/M production after Spike peptide stimulation. K.) Heatmap of data in J.

**Table S1: Antibodies used in study**

| **Antibody** | **Source** | **Clone** | **Identifier (Cat. No.)** |
| --- | --- | --- | --- |
| CD3-BUV395 | BD Biosciences | SK7 | 564001 |
| CD38-BUV496 | BD Biosciences | HIT2 | 612947 |
| CD137-APC | Biolegend | 4-1BB | 309810 |
| CCR7-BUV737 | BD Biosciences | 3D12 | 741786 |
| CD27-BUV805 | BD Biosciences | L125 | 748704 |
| CD8-V500 | BD Biosciences | SK1 | 561618 |
| CD45RA-BV570 | Biolegend | HI100 | 304132 |
| CD45RO-BV605 | Biolegend | UCHL1 | 304238 |
| CD4-BV711 | BD Biosciences | SK3 | 563028 |
| HLA-DR-BV480 | BD Biosciences | G46-6 | 566113 |
| CXCR5-PE-Dazzle 594 | Biolegend | J252D4 | 356928 |
| ICOS-AF700 | Biolegend | C398.4A | 313528 |
| CD69-BV650 | Biolegend | FN50 | 310934 |
| CD134-APC-Fire 750 | Biolegend | Ber-ACT35 | 350032 |
| CD25-BV785 | Biolegend | BC96 | 302638 |
| CXCR3-PE | Biolegend | G025H7 | 353706 |
| Granzyme A-AF700 | Biolegend | CB9 | 507210 |
| Granzyme B-PE-Cy7 | Biolegend | QA18A28 | 396410 |
| IL-21-PE | Biolegend | 3A3-N2 | 513004 |
| Granzyme M-AF488 | Invitrogen | 4B2G4 | 53-9774-42 |
| Perforin-APC | Biolegend | B-D48 | 353312 |
| TNF-α-BV785 | Biolegend | MAb11 | 502948 |
| IL-6-BV421 | BD Biosciences | MQ2-13A5 | 536279 |
| PD-1-BV421 | BD Biosciences | EH12.1 | 562516 |

**Table S2: Related to Fig. 2A-C, G-H**

| **Patient Demographics** | **NP** | **CC** | **HC** |
| --- | --- | --- | --- |
| Participants | 40 | 37 | 22 |
| Sex | 27F, 13M | 18F, 19M | 11F, 11M |
| Age (range) | 48.3 (25-74) | 41.6 (22-73) | 34.82 (22-66) |
| Race/Ethnicity | 29 White  3 Asian  2 Black  5 Hispanic | 20 White  8 Asian  1 Black  8 Hispanic | 10 White  9 Asian  1 Hispanic  1 Other |
| Vaccinated (%; N-M peptides only) | 0 (0) | 15 (40.5) | 2 (9.1) |
| Comorbidities (%) | 26 (65.0) | 11 (29.7) | 8 (36.3) |
| Autoimmune | 10 (25) | 2 (5.4) | 3 (13.6) |
| Cardiac | 12 (30) | 5 (13.5) | 1 (4.5) |
| Respiratory | 9 (22.5) | 2 (5.4) | 2 (9.1) |
| Metabolic | 9 (22.5) | 5 (13.5) | 3 (13.6) |
| Cancer | 2 (5) | 0 (0) | 1 (4.5) |
| Psychiatric | 2 (5) | 2 (5.4) | 0 (0) |
| Neurological | 5 (12.5) | 1 (2.7) | 1 (4.5) |
| Drug Abuse | 5 (12.5) | 0 (0) | 0 (0) |
| Other | 0 (0) | 0 (0) | 0 (0) |
| Hospitalization (%) | 9 (22.5) | 1 (2.7) | N/A |
| Mild Illness (%) | 70 | 97.3 | N/A |
| Days post-symptom onset (range) | 182.5  (20-407) | 162.4  (39-787) | N/A |
| Neurologic symptoms (%) | 40 (100) | 0 (0) | 0 (0) |
| Brain Fog | 37 (92.5) |  |  |
| Fatigue | 34 (85) |  |  |
| Headache | 29 (72.5) |  |  |
| Dizziness | 20 (50) |  |  |
| Anosmia | 22 (55) |  |  |
| Dysgeusia | 23 (57.5) |  |  |
| Myalgias | 24 (60) |  |  |

**Table S3: Related to Figure 2D**

| **Patient Demographics** | **NP** | **CC** |
| --- | --- | --- |
| Participants | 25 | 17 |
| Sex | 15F, 10M | 11F, 6M |
| Age (range) | 47.7 (30-73) | 34.2 (22-70) |
| Race/Ethnicity | 11 White  3 Asian/Pacific Islander  4 Black  7 Hispanic | 11 White  4 Asian/Pacific Islander  2 Hispanic |
| Comorbidities (%) | 16 (64) | 2 (11.8) |
| Autoimmune | 2 (8) | 0 (0) |
| Cardiac | 5 (20) | 1 (5.9) |
| Respiratory | 6 (24) | 1 (5.9) |
| Metabolic | 10 (40) | 1 (5.9) |
| Cancer | 1 (4) | 0 (0) |
| Psychiatric | 2 (8) | 1 (5.9) |
| Neurological | 5 (20) | 0 (0) |
| Drug Abuse | 2 (8) | 0 (0) |
| Other | 0 (0) | 0 (0) |
| Vaccinated (%) | 20 (80)  15 (60) Pfizer  3 (12) Moderna  2 (8) Janssen | 15 (88.2)  12 (70.5) Pfizer  3 (17.6) Moderna |
| Days post-symptom onset  (range) | 263.9  (64-682) | 260.4  (57-616) |
| Neurologic symptoms (%) | 25 (100) | 0 (0) |
| Brain Fog | 22 (56.4) |  |
| Fatigue | 21 (53.8) |  |
| Headache | 20 (51.3) |  |
| Dizziness | 17 (43.6) |  |
| Anosmia | 18 (46.2) |  |
| Dysgeusia | 18 (46.2) |  |
| Myalgias | 17 (43.6) |  |

**Table S4: Related to Fig. 3**

| **Patient Demographics** | **NP** | **CC** | **HC** |
| --- | --- | --- | --- |
| Participants | 41 | 24 | 12 |
| Sex | 32F, 9M | 12F, 12M | 8F, 4M |
| Age (range) | 47.0 (18-83) | 42.3 (23-73) | 37.2 (22-66) |
| Race/Ethnicity | 24 White  6 Black  1 Asian  10 Hispanic | 12 White  1 Black  3 Asian  8 Hispanic | 3 White  1 Black  3 Asian  2 Hispanic  1 Other |
| Vaccinated (%; N-M peptides only) | 0 (0) | 10 (41.7) | 2 (16.7) |
| Comorbidities (%) | 18 (69.2) | 8 (33.3) | 5 (41.7) |
| Autoimmune | 7 (17.1) | 0 (0) | 1 (8.3) |
| Cardiac | 14 (34.1) | 1 (7.1) | 1 (8.3) |
| Respiratory | 7 (17.1) | 0 (0) | 2 (16.7) |
| Metabolic | 13 (31.7) | 1 (7.1) | 3 (25) |
| Cancer | 1 (2.4) | 0 (0) | 1 (8.3) |
| Psychiatric | 2 (4.9) | 2 (14.3) | 0 (0) |
| Neurological | 6 (14.6) | 0 (0) | 0 (0) |
| Drug Abuse | 1 (2.4) | 0 (0) | 0 (0) |
| Other | 2 (4.9) | 0 (0) | 0 (0) |
| Hospitalization (%) | 8 (19.5) | 0 (0) | NA |
| Mild Illness (%) | 31 (75.6) | 24 (100) | NA |
| Days post-symptom onset  (range) | 231.55  (64-580) | 213.1  (39-787) | NA |
| Neurologic symptoms (%) | 41 (100) | 0 (0) | 0 (0) |
| Brain Fog | 37 (90.2) |  |  |
| Fatigue | 36 (87.8) |  |  |
| Headache | 27 (65.8) |  |  |
| Dizziness | 22 (53.7) |  |  |
| Anosmia | 22 (53.7) |  |  |
| Dysgeusia | 20 (48.8) |  |  |
| Myalgias | 24 (58.5) |  |  |

**Table S5: Related to Fig. 4**

| **Patient Demographics** | **NP** | **CC** | **HC** |
| --- | --- | --- | --- |
| Participants | 62 | 29 | 16 |
| Sex | 40F, 22M | 16F, 13M | 10F, 6M |
| Age (range) | 47.5 (18-70) | 41.1 (19-73) | 34.3 (22-66) |
| Race/Ethnicity | 40 White  5 Asian  6 Black  11 Hispanic | 14 White  6 Asian  1 Black  8 Hispanic | 8 White  6 Asian  1 Black  6 Hispanic  1 Other |
| Vaccinated (%; N-M peptides only) | 2 (3.2) | 9 (31.0) |  |
| Comorbidities (%) | 44 (67.7) | 8 (27.5) | 6 (27.2) |
| Autoimmune | 16 (25.8) | 1 (3.4) | 2 (9.1) |
| Cardiac | 17 (27.4) | 4 (13.8) | 0 (0) |
| Respiratory | 14 (22.6) | 1 (3.4) | 0 (0) |
| Metabolic | 18 (29.0) | 3 (10.3) | 2 (9.1) |
| Cancer | 1 (1.6) | 0 (0) | 0 (0) |
| Psychiatric | 3 (4.8) | 2 (6.9) | 0 (0) |
| Neurological | 9 (14.5) | 0 (0) | 0 (0) |
| Drug Abuse | 1 (1.6) | 0 (0) | 0 (0) |
| Other | 3 (4.8) | 2 (6.9) | 2 (9.1) |
| Days post-symptom onset  (range) | 209.5 | 190.8 | NA |
| Neurologic symptoms (%) | 62 (100) | 0 (0) | 0 (0) |
| Brain Fog | 53 (85.5) |  |  |
| Fatigue | 51 (82.3) |  |  |
| Headache | 42 (67.7) |  |  |
| Dizziness | 33 (53.2) |  |  |
| Anosmia | 35 (56.5) |  |  |
| Dysgeusia | 35 (56.5) |  |  |
| Myalgias | 42 (67.7) |  |  |

**Table S5: Related to Fig.6**

| **Patient Demographics** | **NP** | **CC** |
| --- | --- | --- |
| Participants | 48 | 20 |
| Sex | 33F, 15M | 14F, 6M |
| Age (range) | 44.2 (19-83) | 38.8 (22-70) |
| Race/Ethnicity | 33 White  2 Asian  4 Black  9 Hispanic | 12 White  3 Asian  5 Hispanic |
| Vaccinated (%) | 0 (0) | 0 (0) |
| Comorbidities (%) | 34 (70.8) | 4 (25.0) |
| Autoimmune | 8 (16.7) | 1 (5.0) |
| Cardiac | 10 (20.8) | 2(10.0) |
| Respiratory | 10 (20.8) | 1 (5.0) |
| Metabolic | 12 (25.0) | 2 (10.0) |
| Cancer | 2 (4.2) | 0 (0) |
| Psychiatric | 1 (2.1) | 2 (10.0) |
| Neurological | 4 (8.3) | 0 (0) |
| Drug Abuse | 1 (2.1) | 0 (0) |
| Other | 8 (16.7) | 0 (0) |
| Days post-symptom onset  (range) | 214.0 | 207.1 |
| Neurologic symptoms (%) | 48 (100) | 0 (0) |

**Table S6: Proteins associated with the enriched Reactome term “Immunoregulatory interactions between a lymphoid and non-lymphoid cell”**

| **Protein** | **Enriched in NP or CC** | **Running Enrichment Score (ES)** |
| --- | --- | --- |
| KLRC1 | NP | -0.39239 |
| CD200 | NP | -0.38915 |
| CDH1 | NP | -0.37979 |
| KIR2DL3 | NP | -0.37263 |
| KIR2DS2 | NP | -0.36381 |
| LILRA4 | NP | -0.35871 |
| CD3E | NP | -0.35115 |
| ITGB2 | NP | -0.34409 |
| KIR2DL4 | NP | -0.33415 |
| KLRB1 | NP | -0.32435 |
| PVR | NP | -0.31892 |
| SIGLEC9 | NP | -0.31746 |
| HLA-E | NP | -0.30916 |
| CD247 | NP | -0.30232 |
| LILRA6 | NP | -0.29224 |
| HLA-C | NP | -0.28728 |
| SLAMF6 | NP | -0.29272 |
| NCR3LG1 | NP | -0.29128 |
| FCGR2B | NP | -0.28062 |
| LILRB1 | NP | -0.27737 |
| LILRA1 | NP | -0.2648 |
| ICAM1 | NP | -0.25376 |
| HLA-G | NP | -0.24127 |
| JAML | NP | -0.22953 |
| ITGB7 | NP | -0.21957 |
| SIGLEC12 | NP | -0.21639 |
| CD300LG | NP | -0.22525 |
| PIANP | NP | -0.2135 |
| CRTAM | NP | -0.20012 |
| LILRA5 | NP | -0.18553 |
| C3 | NP | -0.17163 |
| LILRB5 | NP | -0.1558 |
| CD300A | NP | -0.14271 |
| CD3G | NP | -0.13346 |
| TREM2 | NP | -0.11734 |
| NPDC1 | NP | -0.10121 |
| NCR1 | NP | -0.08749 |
| MICA | NP | -0.07991 |
| CD300C | NP | -0.06067 |
| SIGLEC7 | NP | -0.0406 |
| LILRB2 | NP | -0.01876 |
| CD33 | NP | 0.002247 |
| SFTPD | Not enriched | 0.01868 |
| SH2D1A | Not enriched | 0.015075 |
| CD22 | Not enriched | 0.03214 |
| ULBP3 | Not enriched | 0.001475 |
| TREML1 | Not enriched | 0.014966 |
| CXADR | Not enriched | -0.0285 |
| CD226 | Not enriched | -0.03215 |
| CD160 | Not enriched | -0.04767 |
| FCGR1A | Not enriched | -0.0652 |
| ULBP1 | Not enriched | -0.0598 |
| CD40LG | Not enriched | -0.06552 |
| ICAM3 | Not enriched | -0.06051 |
| KIR2DL1 | Not enriched | -0.06609 |
| LILRB4 | Not enriched | -0.06568 |
| SIGLEC6 | Not enriched | -0.07799 |
| MADCAM1 | Not enriched | -0.08978 |
| COL2A1 | Not enriched | -0.11486 |
| SIGLEC5 | Not enriched | -0.13561 |
| LILRA2 | Not enriched | -0.14511 |
| TREM1 | Not enriched | -0.15509 |
| NCR2 | Not enriched | -0.15552 |
| ICAM2 | Not enriched | -0.15561 |
| ITGB1 | Not enriched | -0.15529 |
| CLEC2B | Not enriched | -0.1561 |
| CD1D | Not enriched | -0.15852 |
| CLEC2D | Not enriched | -0.1776 |
| SELL | Not enriched | -0.18834 |
| IGKV1-5 | Not enriched | -0.18705 |
| SIGLEC11 | Not enriched | -0.23738 |
| ICAM5 | Not enriched | -0.24237 |
| CLEC4G | Not enriched | -0.24378 |
| CD1A | Not enriched | -0.24856 |
| OSCAR | Not enriched | -0.25863 |
| SLAMF7 | Not enriched | -0.25763 |
| CD34 | Not enriched | -0.27205 |
| COLEC12 | Not enriched | -0.29594 |
| CD81 | Not enriched | -0.29974 |
| ICAM4 | Not enriched | -0.29917 |
| NECTIN2 | Not enriched | -0.29856 |
| LAIR1 | Not enriched | -0.32821 |

**Table S7: Proteins associated with the enriched term “TASOR target genes”**

| **Protein** | **Enriched in NP or CC** | **Running Enrichment Score (ES)** |
| --- | --- | --- |
| H2BC21 | CC | 0.041461 |
| H2AC11 | CC | 0.082812 |
| H2BC12 | CC | 0.122175 |
| H1-10 | CC | 0.153369 |
| H1-2 | CC | 0.17389 |
| TCEA1 | CC | 0.196269 |
| EIF4B | CC | 0.220251 |
| TPT1 | CC | 0.238227 |
| METTL3 | CC | 0.239878 |
| MAP4K1 | CC | 0.264646 |
| ASNS | CC | 0.284889 |
| PBRM1 | CC | 0.297373 |
| REXO2 | CC | 0.29919 |
| HAT1 | CC | 0.321268 |
| MBD4 | CC | 0.315399 |
| GCLM | CC | 0.334676 |
| WFDC3 | CC | 0.344996 |
| BIRC2 | CC | 0.341277 |
| CYCS | CC | 0.34924 |
| TXNL1 | CC | 0.355495 |
| KIFBP | CC | 0.358501 |
| DDI2 | CC | 0.357891 |
| PRKAR1B | CC | 0.369925 |
| NUDT9 | CC | 0.384326 |
| LSM4 | CC | 0.391249 |
| DPY30 | CC | 0.404312 |
| NDE1 | CC | 0.413568 |
| SELENOF | CC | 0.4194 |
| KIAA2013 | CC | 0.413003 |
| DNAJC19 | CC | 0.421415 |
| BOLA1 | CC | 0.432553 |
| CHEK1 | CC | 0.437323 |
| PCK2 | CC | 0.446057 |
| CNPY4 | CC | 0.448213 |
| CSNK1G2 | CC | 0.450206 |
| CHAC1 | CC | 0.453022 |
| EED | CC | 0.461298 |
| HS2ST1 | Not Enriched | 0.111799 |
| EEF1G | Not Enriched | 0.09529 |
| AHCY | Not Enriched | 0.055954 |
| PYCR1 | Not Enriched | 0.05342 |
| MCTS1 | Not Enriched | 0.049315 |
| MPG | Not Enriched | 0.045742 |
| P2RX6 | Not Enriched | 0.036139 |
| BET1L | Not Enriched | 0.031921 |
| PZP | Not Enriched | 0.014856 |
| TPD52L1 | Not Enriched | 0.005694 |
| CYB5D2 | Not Enriched | 0.005683 |
| GOLGA7 | Not Enriched | 0.005451 |
| TIMM23 | Not Enriched | 0.004839 |
| YARS1 | Not Enriched | -0.00301 |
| DLD | Not Enriched | -0.0036 |
| DPH5 | Not Enriched | -0.00892 |
| VAMP8 | Not Enriched | -0.02547 |
| RIC8A | Not Enriched | -0.02935 |
| CDK2AP2 | Not Enriched | -0.04083 |
| MPC1 | Not Enriched | -0.04306 |
| DLAT | Not Enriched | -0.04384 |
| DDIT4 | Not Enriched | -0.04857 |
| AXIN2 | Not Enriched | -0.05425 |
| SLC3A2 | Not Enriched | -0.05462 |
| ANKRA2 | Not Enriched | -0.05494 |
| NFKBIA | Not Enriched | -0.07652 |
| BAG5 | Not Enriched | -0.07701 |
| IARS1 | Not Enriched | -0.09701 |
| PSMB6 | Not Enriched | -0.10155 |
| GUSB | Not Enriched | -0.12449 |
| HAX1 | Not Enriched | -0.12674 |
| ANTKMT | Not Enriched | -0.12699 |
| EMC8 | Not Enriched | -0.14414 |
| SENP8 | Not Enriched | -0.15367 |
| MRFAP1L1 | Not Enriched | -0.15727 |
| TRAPPC5 | Not Enriched | -0.16972 |
| PKN2 | Not Enriched | -0.17409 |
| C4orf36 | Not Enriched | -0.18719 |
| CDKN2C | Not Enriched | -0.18783 |
| MZF1 | Not Enriched | -0.23681 |
| NEURL4 | Not Enriched | -0.23844 |

**Table S8: Proteins associated with the enriched Reactome term “Interleukin 12 family signaling”**

| **Protein** | **Enriched in NP or CC** | **Running Enrichment Score (ES)** |
| --- | --- | --- |
| ANXA2 | CC | 0.043231 |
| HNRNPF | CC | 0.082275 |
| HNRNPDL | CC | 0.120979 |
| HNRNPA2B1 | CC | 0.156849 |
| PPIA | CC | 0.194463 |
| SOD1 | CC | 0.205477 |
| CFL1 | CC | 0.220405 |
| SNRPA1 | CC | 0.245565 |
| MSN | CC | 0.274722 |
| CA1 | CC | 0.29366 |
| RALA | CC | 0.318667 |
| ARF1 | CC | 0.344132 |
| AIP | CC | 0.366265 |
| IFNG | CC | 0.392739 |
| JAK2 | CC | 0.401636 |
| CNN2 | CC | 0.424036 |
| TYK2 | CC | 0.429644 |
| CAPZA1 | CC | 0.448424 |
| IL12RB1 | CC | 0.460917 |
| GSTO1 | CC | 0.482871 |
| LMNB1 | CC | 0.482785 |
| TCP1 | CC | 0.50295 |
| PSME2 | CC | 0.509627 |
| PITPNA | CC | 0.507715 |
| VAMP7 | CC | 0.487042 |
| STAT1 | CC | 0.498079 |
| MIF | CC | 0.508265 |
| STAT3 | CC | 0.511957 |
| CDC42 | CC | 0.512218 |
| IL23R | Not Enriched | 0.372513 |
| BOLA2 | Not Enriched | 0.358263 |
| P4HB | Not Enriched | 0.345793 |
| SERPINB2 | Not Enriched | 0.310886 |
| SOD2 | Not Enriched | 0.26441 |
| CANX | Not Enriched | 0.20159 |
| HSPA9 | Not Enriched | 0.181974 |
| EBI3 | Not Enriched | 0.164821 |
| PDCD4 | Not Enriched | 0.164713 |
| LCP1 | Not Enriched | 0.15046 |
| IL12RB2 | Not Enriched | 0.14816 |
| MTAP | Not Enriched | 0.116181 |
| IL10 | Not Enriched | 0.100209 |
| TALDO1 | Not Enriched | 0.074589 |
| CRLF1 | Not Enriched | 0.088354 |
| IL27RA | Not Enriched | 0.070053 |
| IL12B | Not Enriched | -0.01133 |
| IL6ST | Not Enriched | -0.00715 |
| GSTA2 | Not Enriched | 0.02272 |

**Table S9: Proteins associated with the enriched WP term “IL3 signaling pathway”**

| **Protein** | **Enriched in NP or CC** | **Running Enrichment Score (ES)** |
| --- | --- | --- |
| YWHAQ | CC | 0.02818 |
| BCL2L1 | CC | 0.068464 |
| TGFB1 | CC | 0.105724 |
| YWHAB | CC | 0.135659 |
| GRB2 | CC | 0.174525 |
| CRKL | CC | 0.212676 |
| BCL2 | CC | 0.2452 |
| SHC1 | CC | 0.273374 |
| BAD | CC | 0.292398 |
| VAV1 | CC | 0.318199 |
| INPP5D | CC | 0.343687 |
| PTPN11 | CC | 0.374893 |
| ENPP3 | CC | 0.398513 |
| JAK2 | CC | 0.425061 |
| PRKACA | CC | 0.452832 |
| SYK | CC | 0.456314 |
| HRAS | CC | 0.47488 |
| PTPN6 | CC | 0.49723 |
| FYN | CC | 0.517706 |
| CXCL8 | CC | 0.530002 |
| CBL | CC | 0.527047 |
| PIK3R1 | CC | 0.531617 |
| LYN | CC | 0.52645 |
| STAT5A | CC | 0.527008 |
| STAT3 | CC | 0.538662 |
| MAPK3 | CC | 0.550775 |
| AKT1 | Not Enriched | 0.53079 |
| SRC | Not Enriched | 0.488154 |
| JUN | Not Enriched | 0.495562 |
| IL3 | Not Enriched | 0.429787 |
| CD69 | Not Enriched | 0.380181 |
| STAT5B | Not Enriched | 0.37043 |
| MAPK1 | Not Enriched | 0.344902 |
| MAP2K1 | Not Enriched | 0.331477 |
| RAPGEF1 | Not Enriched | 0.314284 |
| HCK | Not Enriched | 0.315327 |
| CSF2RB | Not Enriched | 0.292595 |
| RAF1 | Not Enriched | 0.251805 |
| MAPK8 | Not Enriched | 0.257686 |
| IL5RA | Not Enriched | 0.158637 |
| SOS1 | Not Enriched | 0.035535 |
| CD86 | Not Enriched | -0.02642 |
| IL3RA | Not Enriched | 0.011907 |
